# Validation of methods for forecasting the frequency of non-vaccine serotypes after introduction or switch of a pneumococcal conjugate vaccine

**DOI:** 10.64898/2026.04.16.26351051

**Authors:** Deus Thindwa, Daniel M Weinberger

## Abstract

**Background:** To anticipate the impact of new pneumococcal vaccines and guide future updates, accurate forecasts of changes in non-vaccine serotypes (NVTs) are needed. We developed and evaluated three models that incorporated different assumptions about the way in which NVTs will increase and generated ensemble predictions for the frequency of NVTs in different post-pneumococcal conjugate vaccines (PCV) periods.

**Methods:** We analyzed age- and serotype-specific invasive pneumococcal disease (IPD) cases from the United States CDC’s Active Bacterial Core surveillance during the pre-PCV (1998-1999), early post-PCV7 (2000-2004), late post-PCV7/pre-PCV13 (2005-2009), early post-PCV13 (2010-2014), and late post-PCV13 (2015-2019) periods. These data were augmented with IPD cases from several countries and combined with serotype-specific invasiveness to infer serotype-specific carriage prevalence. Three models (“Ranking”, “Proportionate”, “NFDS-lite”) generated independent predictions of post-PCV IPD frequencies, which were integrated using an accuracy-weighted ensemble. Model performance was evaluated using the normalized root mean square error (NRMSE).

**Results:** A total of 23,959 non-PCV7 and 15,580 non-PCV13 cases were analyzed. NVT cases increased from the pre-PCV7 to the late post-PCV7 and post-PCV13 periods. The accuracy of predictions across age groups and models was consistent and high during the post-PCV13 periods but varied during the post-PCV7 periods. The Proportionate model (NRMSE=0.70-3.95) outperformed the NFDS-lite (NRMSE=0.93-8.91) and Ranking (NRMSE=1.51-5.37) models during the early-post-PCV7 period, whereas the NFDS-lite model (NRMSE=1.55-9.82) was superior to the Proportionate (NRMSE=1.45-10.22) and Ranking (NRMSE=1.86-11.35) models during the late post-PCV7 period. The Ensemble model improved on these individual models.

**Conclusions:** The Ensemble model offers a tool for forecasting serotype patterns to inform pneumococcal vaccines impact and future pneumococcal vaccine formulation.

## Introduction

*Streptococcus pneumoniae* (pneumococcus) causes a large burden of invasive pneumococcal disease (IPD), particularly among children under five years of age and adults aged 50 years and older [1]. In the United States (US), the seven-valent pneumococcal conjugate vaccine (PCV7) was introduced among children in 2000 and was later replaced by the 13-valent formulation (PCV13) in 2010 [2]. Routine use of PCV13 was subsequently expanded to all older adults (≥65 years-old) starting in 2014 to complement a polysaccharide vaccine (PPSV23), which had been used since 1984 [3,4]. Recently, the 15-valent (PCV15) and 20-valent (PCV20) vaccines have been recommended for both pediatric and adult populations [5,6]. Several next-generation pneumococcal vaccines including those targeting 24-30+ serotypes, are currently under clinical investigation and are anticipated to be used in children and adults in the near future [7–9].

Although pneumococcal vaccines confer effective protection against disease caused by vaccine-targeted serotypes (VTs) in both vaccinated individuals and in unvaccinated populations [10–12], a considerable burden of disease persists. This burden is largely attributed to the resurgence or persistence of VTs and the replacement of VTs with non-vaccine serotypes (NVTs) [13–15]. Because these dynamics can diminish the overall public health impact of pneumococcal vaccines, it is essential to understand post-vaccination changes in serotype carriage and IPD. This information can be used to guide decisions on use of available vaccines and to guide the selection of serotypes for future pneumococcal vaccines.

Several factors shape predictions of pneumococcal serotype dynamics following the introduction or switch of pneumococcal vaccines. Higher-valency vaccines target a broader spectrum of serotypes that cause severe disease [16]. However, the reduced immunogenicity of shared serotypes in higher-valency vaccines might erode protection [17,18]. After introduction or switch of a pneumococcal vaccine, the rate at which VTs decline and NVTs expand to fill the vacated nasopharyngeal niche varies across settings partly due to differences in the baseline local force of VT infection or in local contact patterns [19–21]. Limited surveillance of IPD cases and serotyping may bias post-vaccine predictions. Moreover, newly included serotypes in higher-valency vaccines may confer cross-protection against existing VTs or NVTs, adding further complexity to predicting NVT dynamics in the post-vaccine period [22].

Pneumococcal colonisation is a precursor for disease. The pneumococcus colonizes the nasopharynx of young children with some serotypes that are more competitive than others [23,24]. This competition has direct implications for serotype trajectories in carriage and subsequent disease after introduction or switch of a pneumococcal vaccine. Model-based studies have previously explored post-PCV trajectories of serotypes or serotype groups for PCV impact or vaccine formulation [25–33]. Most models have largely focused on the period after PCV introduction rather than on a switch from one vaccine to another [28,30,32]. Predicting changes following a switch in vaccines poses additional challenges including accounting for the relative effectiveness of the vaccine against shared serotypes [17,18,34,35]. Some studies have also (a) aggregated serotypes into broader VT and NVT groups without capturing individual serotype behavior [29,31,32], (b) described changes in colonization based on observed post-PCV carriage isolates, which may not be available during forecasting [32], and (c) used carriage data from a limited source location [25].

We address several limitations of previous studies by developing three statistical models that make distinct assumptions about how much NVTs expand and combining these independent predictions into an ensemble. The aim is to generate more robust predictions of NVT IPD frequencies following introduction or switch of a pneumococcal vaccine.

## Methods

### Modeling overview

The goal of this analysis was (a) to use data on serotype-specific IPD case counts during the pre-PCV introduction period (1998-1999) to forecast serotype IPD counts during early (2000-2004) or late (2005-2009) post-PCV7 period, and (b) to use data from the late post-PCV7 (pre-PCV13) period (2005-2009) to forecast serotype IPD counts during the early (2010-2014) or late (2015-2019) post-PCV13 periods. The forecasted serotype-specific IPD case count is a function of the serotype-specific IPD case count during the pre-PCV period (“baseline period”), the relative effectiveness of different PCVs against VT IPD, vaccination coverage, and the expected change in post-PCV serotype carriage.

To predict the change in post-vaccine colonization with specific serotypes, we developed an ensemble comprising three models (Proportionate, Ranking and NFDS-lite), each embodying distinct assumptions about how NVTs increase following the introduction or switch of PCVs. In each model, the baseline serotype-specific count of IPD cases and the serotype-specific invasiveness were used to infer the serotype-specific carriage prevalence in that baseline period. This inferred baseline carriage prevalence was, in turn, used as an input to predict the serotype-specific change in the prevalence of carriage of each serotype (Supplementary Figure 1).

### IPD data sources

Age- and serotype-specific IPD case data from 1998-2019 were sourced from the US Center for Disease Control and Prevention (CDC)’s Active Bacterial Core surveillance (ABCs) project [2]. We used data from the eight surveillance sites that continuously reported data since 1998.

Additional data on serotype-specific IPD rates, which were used to augment US IPD cases during the baseline periods, were obtained from surveillance programs of countries that switched from PCV7 to PCV13, including from the Australian National Notifiable Disease Surveillance System (2009-2019) [36], the UK/English Health Security Agency (2004-2016) [37], and the Global Pneumococcal Sequencing project at the Wellcome Sanger Institute for Israel (2006-2016) and The Gambia (2007-2016) [38]. Further details about the data from these surveillance programs are in the supplement (Supplementary Text 1).

### Data augmentation and uncertainty of baseline IPD cases

Some serotypes were associated with zero or a few mean annual cases of IPD in the US during the baseline period. Zero counts are particularly problematic for our prediction framework, making it impossible to obtain predictions greater than zero. Low case counts are also unstable and associated with considerable uncertainty. Thus, we carried out data augmentation to obtain more robust serotype-specific estimates of IPD rates during the baseline period. We fitted a random-effects Poisson regression model using Integrated Nested Laplace Approximation with US IPD case counts as the outcome variable, serotype as a random effect, log-transformed rates of IPD cases during the pre-PCV periods in Australia, England, Israel, and The Gambia as predictors, and the number of baseline period years of IPD surveillance in the US as an offset. The model was fitted separately for each age group and for each pre-PCV7 or pre-PCV13 period. Uncertainty of the fitted incidence was obtained by sampling from a multivariate normal distribution 10,000 times using the model-fitted values and covariance matrix, summarized through 95% quantiles of samples. Details of model fitting and uncertainty are included in the supplement (Supplementary Text 2, Supplementary Figure 2).

### Serotype invasiveness and uncertainty

We obtained the point estimates and standard deviations of serotype invasiveness from published literature, defined as the number of IPD cases per carrier per year [39]. The invasiveness of a given serotype among children was assumed to be the same across surveillance time and for other age groups. Serotypes without invasiveness estimates were either informed by the mean of invasiveness estimates within their serogroup or by the mean of invasiveness estimates from all serotypes if serogroup estimates were unavailable. Uncertainty of serogroup invasiveness was obtained by sampling from a normal distribution 10,000 times using the mean and standard deviation, summarized through 95% quantiles of samples. Details of invasiveness and uncertainty are included in the supplement (Supplementary Text 3 and Supplementary Figure 3).

### Inferred prevalence of pre-PCV serotype carriage

The statistical models are based on carriage prevalence, which is not measured in our population. Thus, baseline carriage prevalence is inferred based on (modelled) baseline IPD incidence and invasiveness [28,33,39]. To propagate the uncertainty of the estimated serotype-specific carriage prevalence, we combined 10,000 samples of invasiveness and 10,000 samples of baseline IPD case estimates for each serotype.

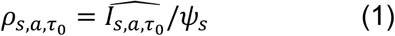

Where ρ*_s,a,τ0_* is the inferred proportion of carriage of serotype (*s*) in an age group (*a*) during the baseline period 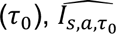 is the fitted serotype IPD cases during the baseline period and ψ_s_ is the serotype invasiveness estimate.

### Forecasting model for IPD

The predicted number of IPD cases for each NVT in an age group in the post-PCV period is a function of the estimated number of IPD cases of that NVT in that age group during the baseline period, the relative vaccine efficacy against IPD of each VT, vaccination coverage, and the change in serotype carriage in that age group as predicted by the model 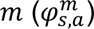 when introducing or switching PCV.

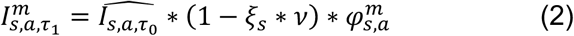

Where 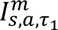 is the predicted number of IPD cases for each serotype (s) in an age group (*a*) during the post-PCV period (τ_1_) informed by a specific model 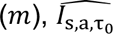 is the estimated number of IPD cases of each serotype in an age group during the baseline period (τ_0_), ψ_s_ is the relative efficacy of the PCV against IPD of each serotype, *v* is the vaccination coverage, and 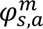 is the predicted change in serotype colonisation in an age group by a model *m* ∈ {Proportionate, Ranking, NFDS-lite}. Details of the relative PCV efficacy against each VT are described in Supplementary Table 1.

### Predicted change in serotype-specific carriage

We considered three models (Proportionate, Ranking, NFDS-lite) that make different assumptions about how the frequency of NVTs will expand in the post-vaccine period.

#### (a) Proportionate model

The simplest assumption is that there is a single constant multiplier applied to the pre-vaccine prevalence of NVTs to predict post-vaccine prevalence of NVTs. The magnitude of this multiplier is determined by how much VTs decrease (how much of a gap in the niche is left to fill). The decrease in VTs is determined by the relative direct effect against carriage for one PCV vs another PCV [35,40]. This model is based on previous studies [31,32], but was generalized to account for the introduction of a pneumococcal vaccine or a switch to another vaccine that differs in effectiveness or serotype coverage. Full details of the Proportionate model are in the supplement (Supplementary Text 4).

#### (b) Ranking model

Despite the shifts in dominant serotypes that occur among colonized individuals after introducing a new pneumococcal vaccine, the frequencies of the top-ranked serotypes are somewhat consistent [19]. For instance, regardless of which specific serotypes are most common, the top serotype has a frequency of p1, the second serotype has a frequency of p_2_, etc. The Ranking model used this pattern to obtain more accurate predictions of the change in frequency for each serotype. Unlike the Proportionate model, this approach determined a serotype-specific expansion factor based on the expected rank and the corresponding prevalence of those ranks in the baseline and post-vaccine periods. We assumed that the ordering of the NVTs was maintained, but the rank shifted based on the removal of VTs. Thus, the Ranking model generated predictions of serotype prevalence ratios based on prevalence rank order of serotypes. Details of the Ranking model are in the supplement (Supplementary Text 5, Supplementary Table 2, Supplementary Figure 4).

#### (c) Negative-frequency-dependent selection ‘lite’ model

The serotypes that increase in frequency following the use of pneumococcal vaccines are often related to those that are included in the vaccine, due to similar biological, genetic and ecological characteristics that allow them to exploit the same nasopharyngeal niche vacated by the VT [41]. For instance, serotype 19A and 6A/C surged when 19F and 6B declined, respectively. In this model, we assumed that the frequency of specific capsular genes was stable following PCV introduction or switch, even if the specific serotypes change. We used the computational approach developed for negative frequency-dependent selection (NFDS) [26,42], but focused only on the genes in the capsular biosynthesis cassette rather than the whole genome (NFDS-lite). This means that NVTs that are genetically related to VTs were predicted to increase the most in carriage. Using our ‘NFDS-lite’ model, we computed the combination of NVTs that best maintained pre-vaccine frequency of capsular genes in the population. Details of the NFDS-lite model are in the supplement (Supplementary Text 6).

### Ensemble model

The ‘Ensemble’ combines serotype predictions from individual models into a single prediction, by assigning weights to each individual prediction [43]. Weights were used dynamically based on past model performance, often to improve accuracy by giving more importance to historically better models or by adjusting for changing patterns, and this leveraged the strengths of each component model. The calculation of weights is given in the supplement (Supplementary Text 7), and the calculation of post-vaccine predictions by the ‘Ensemble’ model is given below,

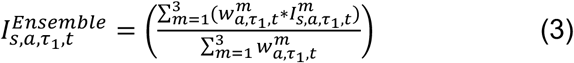

Where 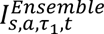 is the Ensemble predicted number of IPD cases at the current time (*t*) for each serotype (s) in a given age group (*a*) during the post-vaccine period 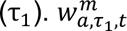 is the weight assigned to the predictions during the current prediction time (*t*) in the post-PCV period (τ_1_) for the given age group and model 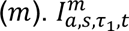 is the predicted number of IPD cases during the post-PCV period of current time for a given serotype in each age group by an individual model.

### Validation of predicted IPD cases

For each component model and the ensemble, we generated 10,000 samples of the mean of the predictions per serotype based on the uncertainty of the carriage prevalence and invasiveness. Then we drew a Poisson sample for each of the 10,000 predictions per serotype to compare to the observed number of IPD cases during the same post-PCV period in the US. We estimated the accuracy between model-predicted and observed IPD cases using the normalized root mean square error (NRMSE) as detailed in the supplement (Supplementary Text 8). The linear and rank relationships between the observed and predicted post-vaccine IPD cases were captured using the Pearson and Spearman correlation coefficients, respectively, with details provided in the supplement (Supplementary Text 9).

### Sensitivity analysis

We conducted a sensitivity analysis by varying the number of years considered for post-vaccine evaluation to assess if the cut-off threshold of early and late post-vaccine years may affect the accuracy of serotype prediction patterns. Thus, we re-ran several model scenarios of ‘early’ and ‘late’ cut-off periods and evaluated their corresponding prediction accuracy. We also lowered the proportion of serotype replacement in the Proportionate model to 5% from the base 95% to assess its effect on the accuracy of post-PCV7 and post-PCV13 serotype predictions.

The R code and US IPD data used in this analysis are freely and publicly available and can be accessed via Github https://github.com/PneumococcalCapsules/serotype_forecasting.

## Results

### Description of the reported serotype IPD data

Serotype data for 39,539 IPD cases caused by NVTs were analyzed. This included 23,959 cases from 1998-2009 [non-PCV7 serotypes], and 15,580 from 2010-2019 [non-PCV13 serotypes]). Cases of NVT increased over time, with the highest recorded cases during the late post-PCV7 period (n=13,417, 33.9%). The greatest number of overall NVT cases occurred among adults aged 50+y (n= 24,744, 62.6%). There were 2,589 (6.6%) cases among <2y and 1,320 (3.3%) among 2-4y children. Dominant NVTs varied across PCV periods with serotypes 14 (9.3%, 17.3%) and 4 (8.5%, 10.0%) during pre-PCV7 and early PCV7 periods, respectively, serotypes 19A (17.9%) and 7F (11.7%) during late PCV7/pre-PCV13 period, serotype 19A (11.0%) and 22F (10.2%) during early post-PCV13 period, and serotype 3 (11.3%) and 22F (10.5%) during late post-PCV13 period (Figure 1). The most common NVTs irrespective of age group and PCV period were 22F (n=4,509, 11.4%), 19A (n=4,025, 10.2%), and 7F (n=2,501, 6.3%), while the least frequent NVTs were serotypes 12B, 15D, and 19C (Figure 1).

**Figure 1.**
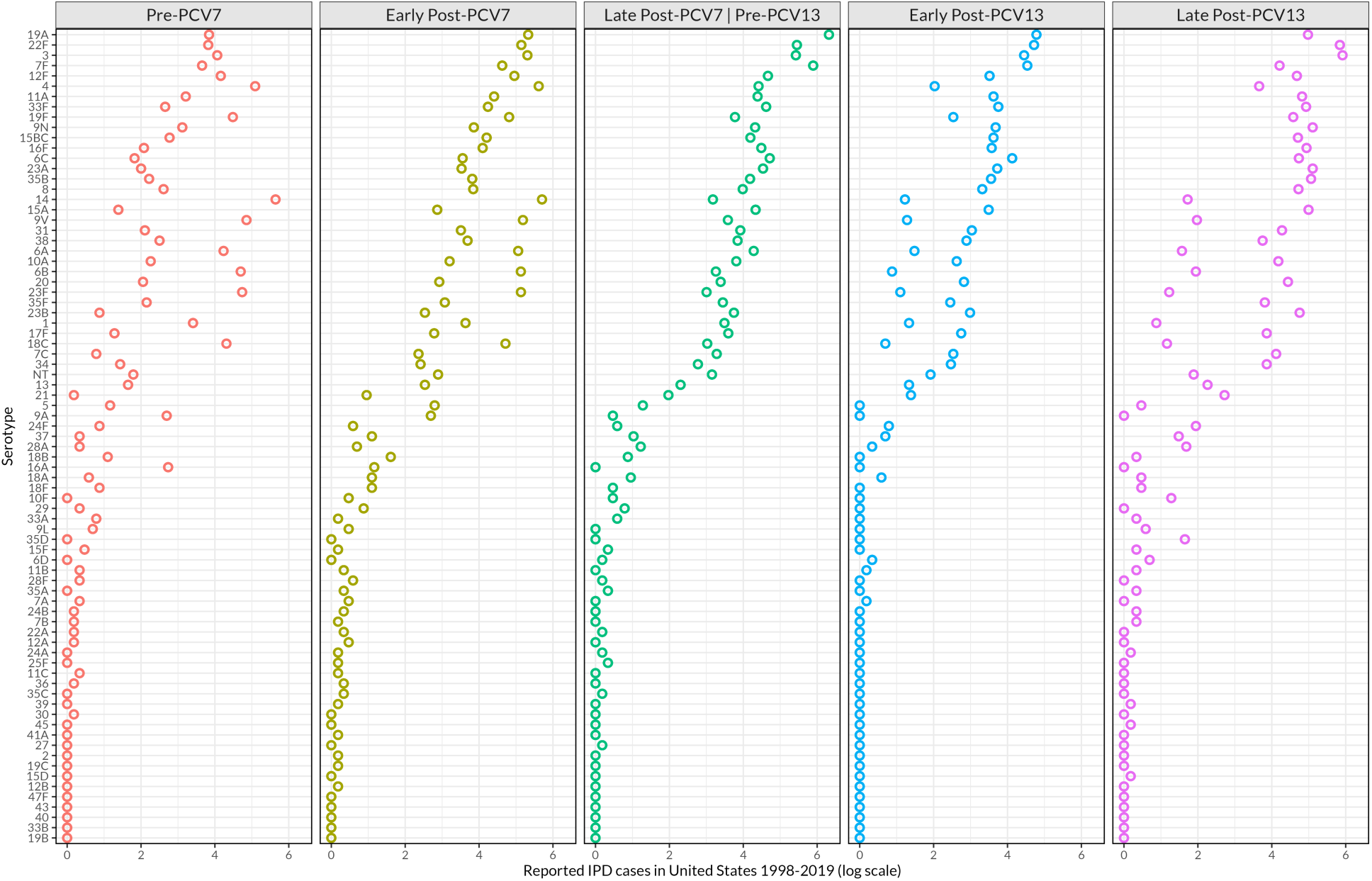
The distribution of serotypes causing invasive pneumococcal disease (IPD) in the United States. The frequency of reported IPD cases (on logarithmic scale) caused by serotypes across all age groups during the considered periods of PCV use including the baseline pre-PCV7 (1998-1999), early post-PCV7 (2000-2004), late post-PCV7 and baseline pre-PCV13 (2005-2009), early post-PCV13 (2010-2014), late post-PCV13 (2015-2019).

### Ensemble model accurately predicts post-vaccine IPD patterns

Overall, out-of-sample ensemble predictions accurately matched the reported IPD cases during the post-PCV7 and post-PCV13 periods. The Spearman and Pearson correlation coefficients were generally higher during the early post-PCV7 compared to the late post-PCV7 but were comparable for the early and late post-PCV13. The accuracy estimates were highest in infants and older adults, and lower in middle aged groups during the post-PCV7 periods. However, the accuracy estimates were comparable across age groups during the post-PCV13 period (Figure 2, Figure 3).

**Figure 2.**
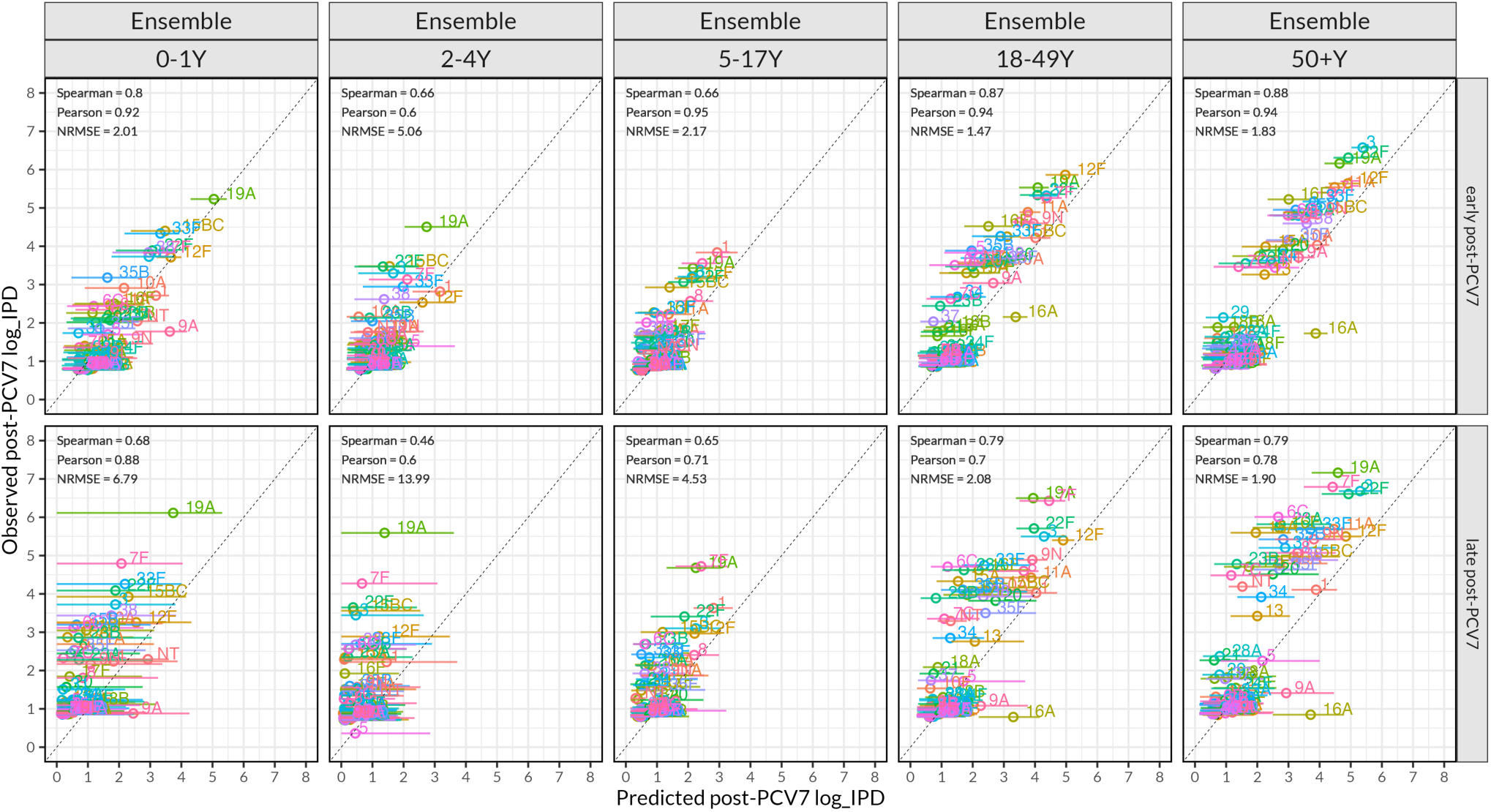
Correlation plot between predicted serotype invasive pneumococcal disease (IPD) cases (x-axis) and observed IPD cases (y-axis) (on log scale), stratified by early post-PCV7 (200-2004) and late post-PCV7 (2005-2009) periods and five age groups (0-1y, 2-4y, 5-17y, 18-49y, and 50+y). Predictions were generated using the Ensemble model, which combined predictions of the three individual models. The colored open dot in the plot represents observed or predicted cases, and the horizontal line through the dot is the 95% confidence interval of the predicted case estimate. The legend embedded within each panel summarizes correlation coefficients and accuracy performance using normalized root mean square error (NRMSE), of the relationship between observed and predicted IPD in the post-PCV periods.

**Figure 3.**
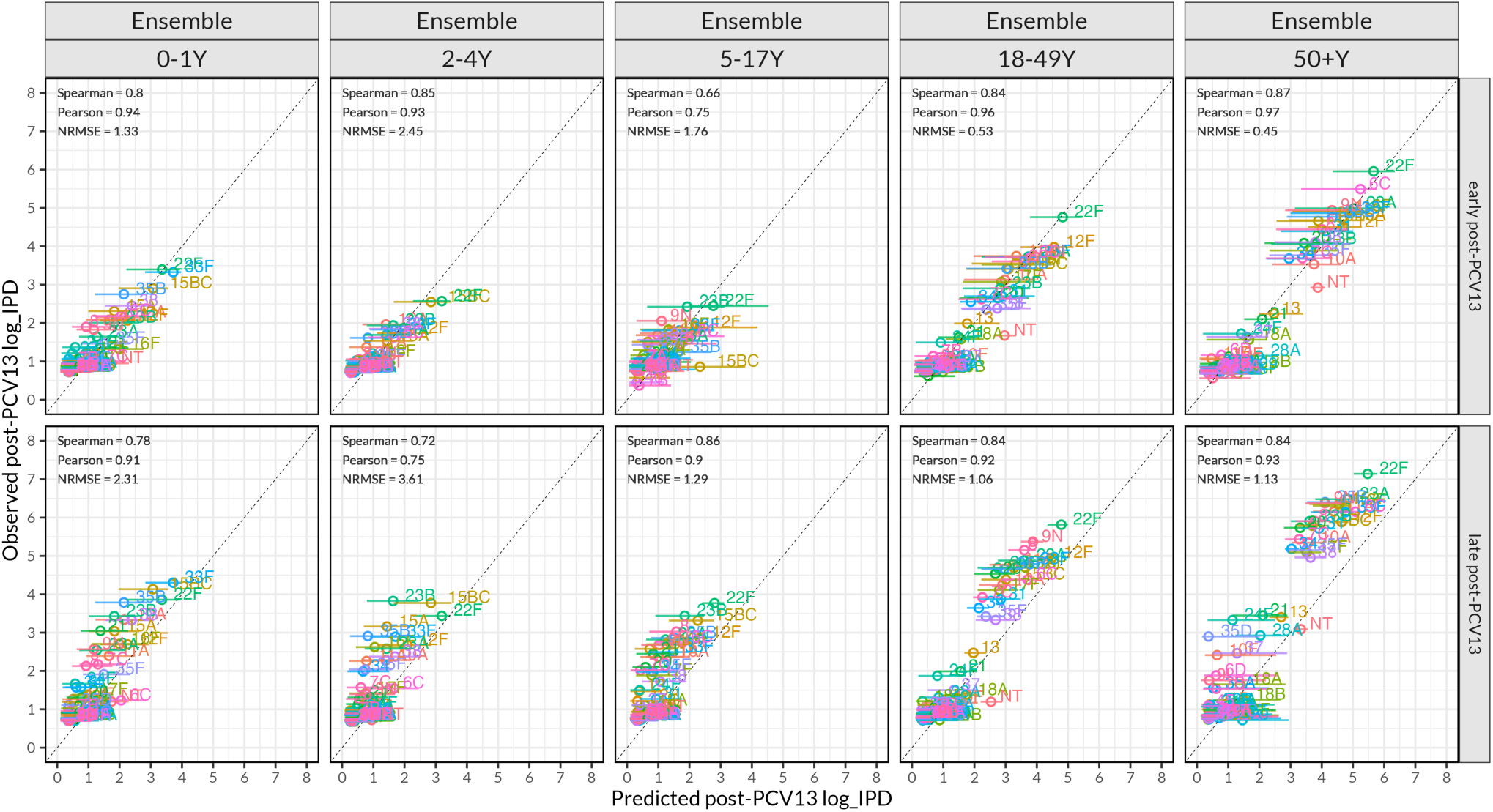
Correlation plot between predicted serotype invasive pneumococcal disease (IPD) cases (x-axis) and observed IPD cases (y-axis) (on log scale), stratified by early post-PCV13 (2010-2014) and late post-PCV13 periods and five age groups (0-1y, 2-4y, 5-17y, 18-49y, and 50+y). Predictions were generated using the Ensemble model, which combined predictions of the three individual models. The colored open dot in the plot represents observed or predicted cases and the horizontal line through the dot is the 95% confidence interval of the predicted case estimate. The legend embedded within each panel summarizes correlation coefficients and accuracy performance using normalized root mean square error (NRMSE), of the relationship between observed and predicted IPD in the post-PCV periods.

### Comparing predictions of serotype patterns between models

Overall, counts of observed serotype IPD cases overlapped with the 95% confidence intervals of predicted cases from at least one individual model. The top-performing individual model varied between time periods and age groups, highlighting the relevance of combining multiple models for more robust predictions. Model predictions were more accurate during the post-PCV13 than post-PCV7 periods. During the late post-PCV7 period, all models slightly underpredicted observed IPD cases of serotypes that caused the most disease. In contrast, serotypes 9A and 16A caused a few cases and were overpredicted by all models during the post-PCV7 period. During the post-PCV13 period, model performances were largely comparable and captured well the observed serotype IPD patterns (Supplementary Figure 5, Supplementary Figure 6, Supplementary Figure 7, Supplementary Figure 8, Supplementary Figure 9, Supplementary Figure 10).

#### (a) Post-PCV7 IPD serotype predictions

During the early post-PCV7 period, the Proportionate model (range: NRMSE = 0.70-3.95) performed best across age groups followed by the NFDS-lite model (NRMSE = 0.93-8.91) and the Ranking model (NRMSE = 1.51-5.37). The Ensemble predictions fell within the predictions of individual models (NRMSE = 0.93-4.06). (Table 1, Supplementary Figure 11). During the late post-PCV7, the NFDS-lite model (NRMSE = 1.55-9.82) largely outperformed other models across age groups, followed by the Proportionate model (NRMSE = 1.45-10.22) and the Ranking model (NRMSE = 1.86-11.35). The Ensemble model synthesized individual model predictions (NRMSE =1.59-10.26) (Table 1, Supplementary Figure 12).

**Table 1.**
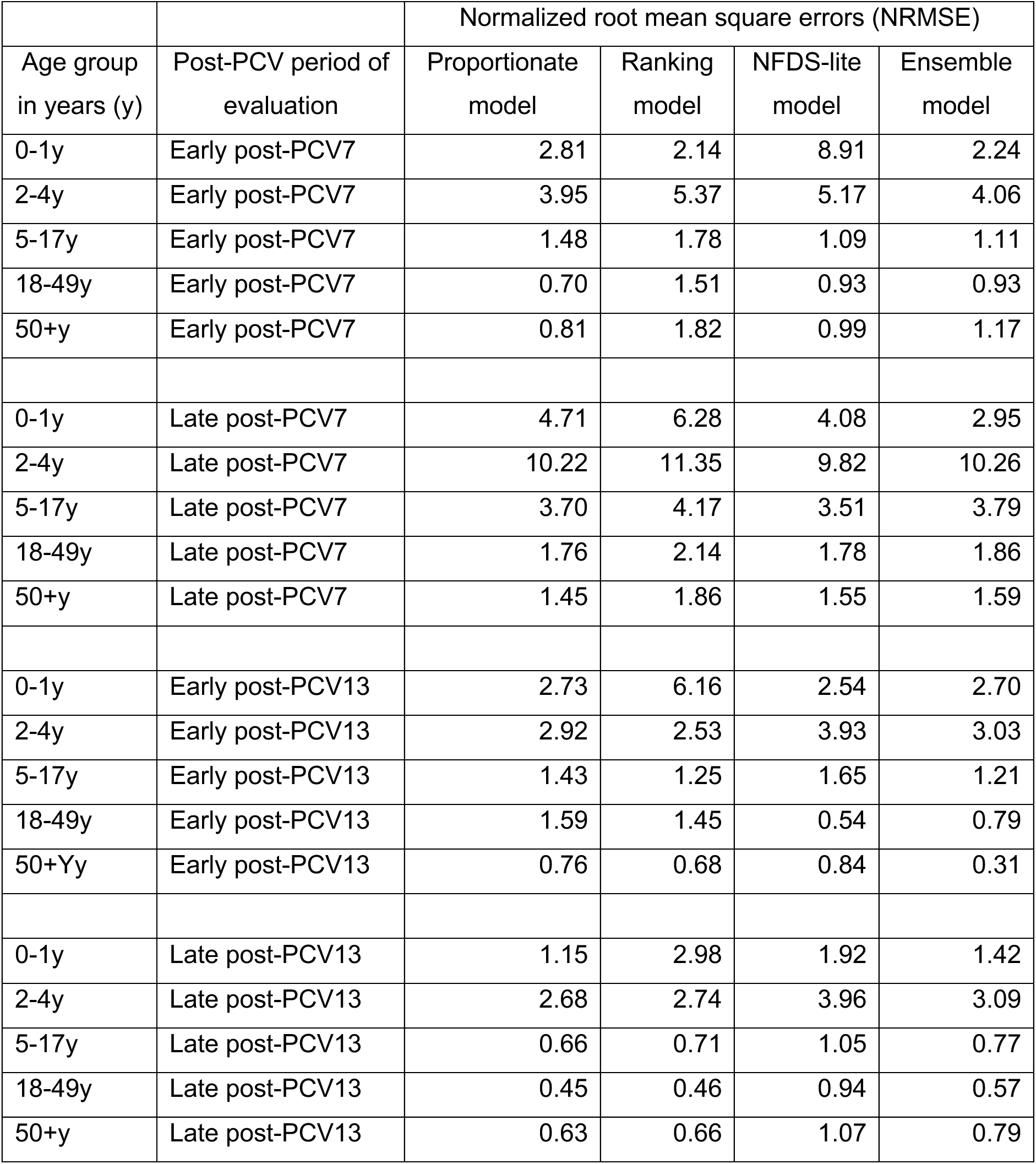
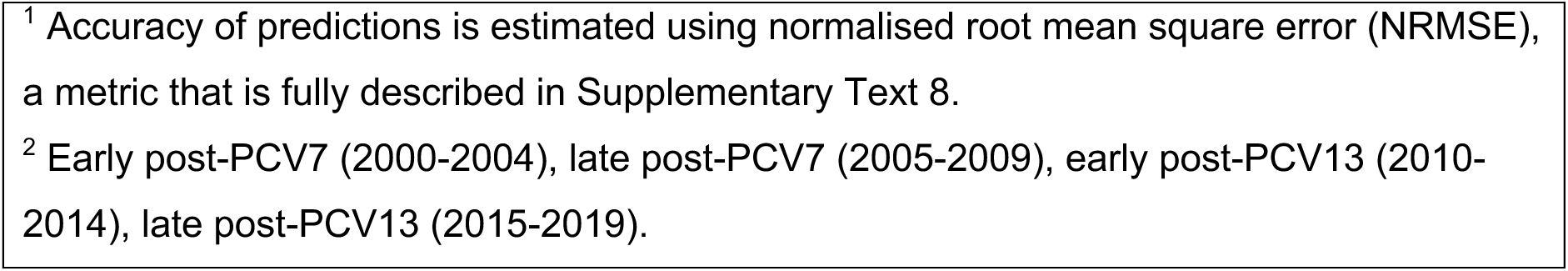
Accuracy of predictions of invasive pneumococcal disease (IPD) cases caused by non-vaccine serotypes and stratified by a five-band age groups, period of post-pneumococcal conjugate vaccine (post-PCV) evaluation and type of the prediction model.

| Age group<br>in years (y) | Post-PCV period of<br>evaluation | Normalized root mean square errors (NRMSE) |  |  |  |
| --- | --- | --- | --- | --- | --- |
|  |  | Proportionate<br>model | Ranking<br>model | NFDS-lite<br>model | Ensemble<br>model |
| 0-1y | Early post-PCV7 | 2.81 | 2.14 | 8.91 | 2.24 |
| 2-4y | Early post-PCV7 | 3.95 | 5.37 | 5.17 | 4.06 |
| 5-17y | Early post-PCV7 | 1.48 | 1.78 | 1.09 | 1.11 |
| 18-49y | Early post-PCV7 | 0.70 | 1.51 | 0.93 | 0.93 |
| 50+y | Early post-PCV7 | 0.81 | 1.82 | 0.99 | 1.17 |
| 0-1y | Late post-PCV7 | 4.71 | 6.28 | 4.08 | 2.95 |
| 2-4y | Late post-PCV7 | 10.22 | 11.35 | 9.82 | 10.26 |
| 5-17y | Late post-PCV7 | 3.70 | 4.17 | 3.51 | 3.79 |
| 18-49y | Late post-PCV7 | 1.76 | 2.14 | 1.78 | 1.86 |
| 50+y | Late post-PCV7 | 1.45 | 1.86 | 1.55 | 1.59 |
| 0-1y | Early post-PCV13 | 2.73 | 6.16 | 2.54 | 2.70 |
| 2-4y | Early post-PCV13 | 2.92 | 2.53 | 3.93 | 3.03 |
| 5-17y | Early post-PCV13 | 1.43 | 1.25 | 1.65 | 1.21 |
| 18-49y | Early post-PCV13 | 1.59 | 1.45 | 0.54 | 0.79 |
| 50+Yy | Early post-PCV13 | 0.76 | 0.68 | 0.84 | 0.31 |
| 0-1y | Late post-PCV13 | 1.15 | 2.98 | 1.92 | 1.42 |
| 2-4y | Late post-PCV13 | 2.68 | 2.74 | 3.96 | 3.09 |
| 5-17y | Late post-PCV13 | 0.66 | 0.71 | 1.05 | 0.77 |
| 18-49y | Late post-PCV13 | 0.45 | 0.46 | 0.94 | 0.57 |
| 50+y | Late post-PCV13 | 0.63 | 0.66 | 1.07 | 0.79 |

<sup>1</sup> Accuracy of predictions is estimated using normalised root mean square error (NRMSE), a metric that is fully described in Supplementary Text 8.
<sup>2</sup> Early post-PCV7 (2000-2004), late post-PCV7 (2005-2009), early post-PCV13 (2010-2014), late post-PCV13 (2015-2019).

#### (b) Post-PCV13 IPD serotype predictions

During the early and late post-PCV13 periods, the Proportionate model (NRMSE = 0.79-2.92 and 0.63-2.68), the Ranking model (NRMSE = 0.68-6.16 and 0.66-2.98) and the NFDS-lite model (NRMSE = 0.54-3.93 and 0.94-3.96) had comparable performances, and their predictions were synthesized by Ensemble predictions (NRMSE = 0.31-3.03 and 0.57-3.09). Unlike the post-PCV7 period, serotypes 9A and 16A were well-predicted in the post-PCV13 period (Table 1, Supplementary Figure 13, Supplementary Figure 14).

### Results of sensitivity analysis

The number of years, following the introduction or switch of a pneumococcal vaccine, needed to predict serotype patterns had an influence on the accuracy of predicted serotype patterns. The prediction accuracy was highest when evaluating accuracy of 4-6 years following PCV7 or PCV13 introduction. However, the prediction accuracy was lowest if the number of evaluation years was less than 4 years or more than 6 years following PCV7 introduction or PCV13 switch (Supplementary Figure 15).

For the base scenario, we assumed that the proportion of serotype replacement was 95% with respect to the proportionate model. Reducing this proportion to 5% resulted in less accurate post-PCV7 serotype predictions. In contrast, this reduction resulted in a slight change in the post-PCV13 serotype predictions, reflecting the distinct speeds of serotype replacement between PCV7 and PCV13 periods (Supplementary Figure 16, Supplementary Figure 17).

Overall, the rank order of NVT predictions was similar regardless of the degree of serotype replacement, highlighting that the main benefit of this modelling approach is in capturing the absolute change in the incidence of the NVTs rather than predicting the rank order of the NVTs.

## Discussion

With new pneumococcal vaccines under development, it is more important than ever to have reliable forecasts for how the NVTs might change following the switch to a vaccine with different serotype composition. We present an ensemble forecasting framework, which includes three methods that each make distinct assumptions about the increasing frequency of NVTs.

Serotype-specific predictions of the frequency of IPD due to NVT matched well with the observed NVT IPD cases, with varying levels of accuracy across the age groups, PCV periods, and individual models. The accuracy of predictions was generally higher during the post-PCV13 than the post-PCV7 periods, although the Ensemble’s effective synthesis controlled for the variances and biases in the individual models. Overall, this study shows individual model strengths while addressing their weaknesses through model combination to improve the post-PCV serotype predictions. Our framework offers a tool for forecasting serotype patterns to inform pneumococcal vaccine impact and future vaccine formulation.

Individual models largely performed well in predicting serotype-specific IPD frequencies. The prediction accuracy of a given model was heterogeneous across age groups and PCV evaluation periods, with the best predictions usually among the infant and older adult groups and in the post-PCV13 period. The higher accuracy of predictions among infants and older adults may relate to populations where VT elimination may have been substantial and led to significant NVT replacement. In contrast, the higher accuracy of post-PCV13 predictions may reflect the use of more robust US surveillance baseline data during the mature PCV7 period than the pre-PCV7 baseline data for predicting post-PCV7 IPD frequencies.

The post-PCV7 prediction accuracies within each age group and PCV period differed across individual models likely due to model assumptions about how the NVTs would increase.

Regardless, the NFDS-lite and Proportionate models usually performed well. Overall, the Ensemble model greatly improved the individual model predictions especially during the post-PCV7 period.

A few NVTs were poorly predicted across models and age groups during the post-PCV7 period e.g., 9A and 16A. Some studies have suggested that the relatedness of 9A and 9V (a serotype targeted by PCV7) could lead to some cross-protection [22]. In the post-PCV13 period, the pre-PCV13 baseline case data corresponding to 9A already reflected any cross-protection from the PCV, unlike the pre-PCV7 baseline data. Other factors may also affect serotype predictions; Invasiveness data are usually available among children but rare among adults who have a limited number of carriage studies [39]. We used invasiveness estimates in children and assumed that other age groups had similar invasiveness. While our assumption was robust given the highly accurate serotype predictions among older adults, this robustness may not be reflective of each serotype.

This study had some limitations: Not all known and unknown potential serotype cross-protection was accounted for. For instance, serotype 6C was categorized as non-PCV13 (NVT) while emerging evidence shows that it may possibly be protected against by PCV13 [44], although not by PCV7 [45]. We inferred carriage prevalence from IPD and invasiveness due to limited temporal carriage data during post-PCV introduction and switch periods in the population.

Colonisation studies are required to inform changes in serotype distribution in different time periods after vaccine switch in order to track untoward serotype behaviour.

Several models have attempted to predict serotype patterns and subsequent PCV impact after PCV introduction [26–29,31,32,42,46]. However, there is a limited body of modeling literature on serotype predictions when switching from a lower to a higher valency pneumococcal vaccine and vice versa, and our methods address such gaps. We developed methods that considered serotype and capsular gene data and factors that have previously not been accounted for in totality including serotype invasiveness and PCVs immunogenicity inferiority [17,18,35]. With newer PCVs being used in children [5,6], and other pneumococcal vaccines under clinical investigation [7–9], our models may help predict the potential impact of these next-generation pneumococcal vaccines before or after a decision to introduce or switch vaccines.

In conclusion, leveraging complementary strengths of different models improved the accuracy of predictions of serotype patterns after PCV introduction or switch. The ensemble model offers a valuable tool for forecasting serotype patterns to inform pneumococcal vaccines impact and future vaccine formulation.

## Acknowledgements

Research reported in this manuscript was fully supported by GSK under award GR128426.

## Author contributions

Conceptualization; DMW

Data curation; DT

Formal analysis; DT, DMW

Funding acquisition; DMW

Investigation; DT, DMW

Methodology; DT, DMW

Project administration; DMW

Resources; DMW

Software; DT

Supervision; DMW

Validation; DT, DMW

Visualization; DT, DMW

Writing - original draft; DT

Writing - review & editing; DT, DMW

All authors read and approved the final manuscript.

## Data availability

The data and code for this analysis are publicly available in the GitHub

https://github.com/PneumococcalCapsules/serotype_forecasting.

