## Supplementary for "Validation of methods for forecasting the frequency of non-vaccine serotypes after introduction or switch of a pneumococcal conjugate vaccine"

**Table of contents**


| | |
|---|---|
| Supplementary Text 1 | PCV timelines of underlying IPD data used in the analysis |
| Supplementary Text 2 | Random effects Poisson regression model for data augmentation |
| Supplementary Text 3 | Invasiveness and sampling from a normal distribution |
| Supplementary Text 4 | Technical details of the Proportionate model |
| Supplementary Text 5 | Technical details of the Ranking model |
| Supplementary Text 6 | Technical details of the NFDS-lite model |
| Supplementary Text 7 | Calculation of the weights for Ensemble serotype predictions |
| Supplementary Text 8 | Calculation of the normalised root mean square error |
| Supplementary Text 9 | Associations between observed and predicted post-PCV IPD cases |
| Supplementary Table 1 | Relative effectiveness of PCVs against each serotype in carriage or in IPD for ‘PCV7’ and ‘PCV13 relative to PCV7’ |
| Supplementary Table 2 | Snapshot demonstration of the Ranking method |
| Supplementary Figure 1 | Modeling overview of forecasting invasive pneumococcal disease (IPD) case frequency caused by non-vaccine serotypes |
| Supplementary Figure 2 | Observed vs predicted IPD cases using a random effects Poisson regression model stratified by PCV periods |
| Supplementary Figure 3 | Distribution of serotype, serogroup and their invasiveness, and their simulations based on normal distribution sampling |
| Supplementary Figure 4 | Serotype prevalence ranks stratified by age groups and PCV periods |
| Supplementary Figure 5 | Comparison of observed and predicted post-PCV7 IPD frequency by age group and post-vaccination period based on Proportionate model. |
| Supplementary Figure 6 | Comparison of observed and predicted post-PCV13 IPD frequency by age group and post-vaccination period based on Proportionate model. |
| Supplementary Figure 7 | Comparison of observed and predicted post-PCV7 IPD frequency by age group and post-vaccination period based on Ranking model. |
| Supplementary Figure 8 | Comparison of observed and predicted post-PCV13 IPD frequency by age group and post-vaccination period based on Ranking model. |
| Supplementary Figure 9 | Comparison of observed and predicted post-PCV7 IPD frequency by age group and post-vaccination period based on NFDS-lite model. |
| Supplementary Figure 10 | Comparison of observed and predicted post-PCV13 IPD frequency by age group and post-vaccination period based on NFDS-lite model. |
| Supplementary Figure 11 | Model comparisons based on predictions of early post-PCV7 serotype invasive pneumococcal disease cases stratified by age groups |
| Supplementary Figure 12 | Model comparisons based on predictions of late post-PCV7 serotype invasive pneumococcal disease cases stratified by age groups |
| Supplementary Figure 13 | Model comparisons based on predictions of early post-PCV13 serotype invasive pneumococcal disease cases stratified by age groups |
| Supplementary Figure 14 | Model comparisons based on predictions of late post-PCV13 serotype invasive pneumococcal disease cases stratified by age groups |
| Supplementary Figure 15 | Model performance in different post-PCV evaluation periods by prediction method |
| Supplementary Figure 16 | Comparison of post-PCV7 predicted and observed IPD based on low or high proportion value of serotype replacement in the Proportionate model |
| Supplementary Figure 17 | Comparison of post-PCV13 predicted and observed IPD based on low or high proportion value of serotype replacement in the Proportionate model |

**Supplementary Text 1**

*PCV timelines of underlying data*

In the US, infant-PCV7 was introduced in July 2000 and replaced by PCV13 in April 2010 under a three-primary dose schedule with a booster (3p+1), with coverage of >80% [1]. In Australia, infant-PCV7 was introduced in January 2005 and replaced by PCV13 in July 2011 under a 3p+0 schedule and coverage of >90%, before switching to a 2p+1 schedule in 2018 [2]. In England/Wales, infant-PCV7 was introduced in September 2006 and replaced by PCV13 in April 2010 under a 2p+1 schedule and coverage of >90% [3]. In Israel, infant-PCV7 was introduced in July 2009 under a 2p+1 schedule then switched to PCV13 in November 2010 with coverage of >80% [4,5]. Lastly, in The Gambia, infant-PCV7 was introduced in August 2009 under a 3p+0 schedule and replaced by PCV13 in July 2011 reaching ~70% coverage [6].

**Supplementary Text 2**

*Random effects Poisson regression model for data augmentation*

We fit a random effects Poisson regression model using Integrated Nested Laplace Approximation (INLA) where indices $i$ and $j$ refer to observation and group. The pre-PCV counts of IPD cases in the United States is the outcome variable ($I_{\tau_0,ij}$), the number of years of reported IPD cases ($Z_{ij}$) is an offset, serotype is a random effect variable ($b_j$), log-transformed rates of IPD cases in Australia ($X_{1,ij}$), England ($X_{2,ij}$), Israel ($X_{3,ij}$) or The Gambia ($X_{4,ij}$) are predictors. Pre-PCV13 IPD rates were available from all countries whereas pre-PCV7 IPD rates were from England, Israel and The Gambia. IPD cases in these countries were considered because, like the United States, they introduced PCV7 and switched to PCV13. The model was fitted separately for each pre-PCV period and age group.

The likelihood:

$$I_{\tau_0,ij} \sim Poisson(\lambda_{ij}) - - - - - (1)$$

Log link with offset

$$\log(\lambda_{ij}) = log(Z_{ij}) + \beta_0 + \beta_1 X_{1,ij} + \beta_2 X_{2,ij} + \beta_3 X_{3,ij} + \beta_4 X_{4,ij} + b_j - - - - - (2)$$

Where $\lambda_{ij}$ is the expected values of the fitted model, $\beta_0$ is the fixed intercept, $\beta_1$, $\beta_2$, $\beta_3$, and $\beta_4$ are coefficients corresponding to predictors $X_1$, $X_2$, $X_3$, and $X_4$.

*Sampling from a multivariate normal distribution*

We generate uncertainty around fitted values of the model above by sampling from a multivariate normal distribution using the estimated model mean and covariance as sampling parameters.

**Supplementary Text 3**

*Invasiveness and sampling from a normal distribution*

Given the estimates of serotype invasiveness ($\psi_s$) and their 95% confidence interval from Løchen et al. [7], we converted these estimates to the log scale ($log\_\psi_s$). We then calculated serotype-specific standard deviation of invasiveness on log scale ($\log\_\omega_s$) using the 95% lower and upper bounds of serotype invasiveness as shown is equation 3. We sample from a normal distribution using mean and standard deviation of invasiveness on the log scale to obtain uncertainty as $\log(X_s) \sim Normal(\log\_\psi_s, \log\_\omega_s)$. Serotypes without invasiveness estimates were either informed by serogroup invasiveness ($\log\_\psi_g$), calculated as the mean of serotype invasiveness within their serogroup $G$ (equation 4) or by mean of serotype invasiveness from all serotypes $N$ if serogroup estimates were unavailable (equation 5).

$$\log\_\omega_s = \frac{([\log(\psi_{s,CI_{0.975}}) - \log(\psi_{s,CI_{0.500}})] + [\log(\psi_{s,CI_{0.500}}) - \log(\psi_{s,CI_{0.025}})])}{(2 * 1.96)} ------ (3)$$

$$\log\_\psi_g = \frac{\sum_s^G \psi_s}{G} ------ (4)$$

$$\log\_\psi_n = \frac{\sum_s^N \psi_s}{N} ------ (5)$$

**Supplementary Text 4**

*Technical details of the Proportionate model*

We developed and expanded this model based on other previous model [8]. This approach assumes that there is a single constant multiplier applied to the pre-PCV prevalence of NVTs to predict post-PCV prevalence of NVTs. The magnitude of this multiplier is determined by how much VTs decrease (how much of a gap in the niche is left to fill). The decrease in VTs is determined by the relative direct effect against carriage for one PCV vs another PCV [9,10]. To infer the change in colonisation, we estimate the size of the gap opened in the nasopharyngeal niche due to the reduction of vaccine serotypes by PCV, and the size of the compensatory increase in non-vaccine serotypes to fill that gap. Thus, the size of the gap is estimated as $G_a = \sum_s(\rho_{s,a,\tau_0} * \zeta_s * \nu * k)$ where $\rho_{s,a,\tau_0}$ is the carriage prevalence of each serotype ($s$) in age group ($a$) during the baseline period ($\tau_0$), $\zeta_s$ is the relative efficacy of PCV against carriage of each serotype, with values closer to 1 indicating no difference between PCVs (Table S1), $\nu$ is the vaccination coverage and $k$ determines if a serotype is targeted by the PCV in the post-PCV period (e.g., $k$ is 1 if vaccine serotypes and 0 otherwise). The total prevalence of non-vaccine serotypes during the baseline period is $C_a = \sum_s(\rho_{s,a,\tau_0} * (1 - k))$. Thus, the expected change in colonisation is $\varphi_a^{Prop} = \left(1 + \frac{\gamma * G_a}{C_a}\right)$ where $\gamma$ is the proportion of the gap filled by non-vaccine serotypes (e.g., $\gamma$=1 complete replacement, $\gamma$=0 no replacement). Thus, $\varphi_a^{Prop}$gives the expected change in carriage prevalence for each NVT following the vaccine introduction/switch.

**Supplementary Text 5**

*Technical details of the Ranking model*

Despite the shifts in dominant serotypes that occur among colonized individuals following introduction of a new PCV, the frequencies of the top serotypes are somewhat consistent [11]. The Ranking model leverages this pattern to get more accurate predictions of the change in frequency for each serotype. Given that the prevalence for serotype $(s)$ in an age group $(a)$, during a baseline period $(\tau_0)$ is $\rho_{s,a,\tau_0}$, then the descending numeric order of serotypes based on prevalence can be denoted as $R_0 = (\{1,2,3,\ldots N\} \mid rank(\rho_{s,a,\tau_0}))$ where $N$ is the total number of serotypes. In a period after PCV introduction or switch $(\tau_1)$, vaccine serotype prevalence is defined as $\rho_{s,a,\tau_1} = \rho_{s,a,\tau_0} * (1 - \zeta_s)$ where $\zeta_s$ is the relative efficacy of PCV against carriage of each serotype (Table S1). The descending numeric order of serotypes during the post-PCV period can be denoted as $R_1 = (\{1,2,3,\ldots N\} \mid rank(\rho_{s,a,\tau_1}))$. Thus, in the post-PCV period, a non-vaccine serotype that caused most carriage among non-vaccine serotypes during the baseline period is assigned a prevalence estimate of a vaccine serotype that caused more carriage overall during the baseline period (preserving rank order e.g., $\rho_{s,a,\tau_1} = (\rho_{s,a,\tau_0} \mid R_1 = R_0)$. The expected change in serotype colonisation during post-PCV is $\varphi_{s,a}^{Rank} = \left(\frac{\rho_{s,a,\tau_0} \mid R_1 = R_0}{\rho_{s,a,\tau_0} \mid R_0}\right)$. Thus, $\varphi_{s,a}^{Rank}$ gives the predicted change in carriage prevalence for each NVT following the vaccine introduction/switch (Table S2).

**Supplementary Text 6**

*Technical details of the Negative Frequency Dependent Selection model variant (NFDS-lite)*

The serotypes that increase in frequency following the use of PCVs are often related to those that were included in the vaccine. In this model, we assume that the frequency of specific capsular genes is stable following the use of a PCV, even if the specific serotypes change. Thus, we use a replicator equation $\frac{d\rho_{s,a,\tau_0}}{d\tau} = \rho_{s,a,\tau_0}(\beta_s - \sum_{s=1}^{N} \rho_{s,a,\tau_0} * \beta_s)$ where $\rho_{s,a,\tau_0}$ is the age group $(a)$ specific frequency of serotype $(s)$ in carriage during baseline period $(\tau_0)$, $d\tau$ is the change in the PCV periods, $\beta_s$ is the fitness of serotype and $(\sum_{s=1}^{N} \rho_{s,a,\tau_0} * \beta_s)$ is the total serotype population fitness in a given age group during baseline period. A vector $\beta$ is the product of matrix $\boldsymbol{K}$ and the vector $(e - f)$ such that $\beta = \boldsymbol{K}(e - f)$. The entry $k_{s,g}$ of the matrix $\boldsymbol{K}$ has values between 0 and 1 to represent the frequency of capsular gene $g$ in serotype $s$, the entry $g^{th}$ of vector $(e - f)$ is to the difference in the frequency of capsular gene $g$ between baseline and post-PCV period. The post-PCV gene frequency $(f_g)$ is considered only for isolates that are of non-vaccine serotype taxa. Thus, the vector $(e - f)$ represents the gap that vaccination creates in pneumococcal population by removing capsular gene loci whereas $\beta_s$ quantifies the ability of a serotype to fill this gap. Based on 167 capsular genes present in 5-80% of taxa in IPD data from the United States, baseline equilibrium frequencies of capsular gene were computed, assuming a negligible effect of recombination on capsular genome over the considered time horizon. We used INLA to estimate $\beta_s$ such that the capsular gene frequencies in post-PCV period closely matched to baseline period. The estimates of $\beta_s$ were multiplied with $\rho_{s,a,\tau_0}$ to obtain predicted prevalence of each serotype. Then the ratio of predicted to observed serotype prevalence $(\varphi_{s,a}^{NFDSl})$ was calculated based on the Ranking method as previously described.

**Supplementary Text 7**

*Calculation of the weights for Ensemble serotype predictions*

An Ensemble combined predictions from individual models into a single prediction, by assigning weights to each individual prediction [12]. The use of weights from a previous prediction period for an ensemble in the current period involves adapting weights dynamically based on past model performance, often to improve accuracy by giving more importance to historically better models or adjusting for changing patterns [13]. Thus, the weighted average of our post-PCV predictions of individual models was computed to generate ensemble predictions to leverage the strength of each component model. Equal weights were assigned to individual models to generate Ensemble predictions during the initial period of early post-PCV7. Weights based on individual model performance during the early post-PCV7 period were used for Ensemble predictions during the late post-PCV7 phase. Likewise, late post-PCV7 individual model performance informed weights for early post-PCV13 Ensemble predictions. And lastly, early post-PCV13 weights informed late post-PCV13 Ensemble predictions. Subsequent weights, after initial equal-weights, were calculated based on normalised root mean square error (NRMSE) scaled between (0,1). The calculations of NRMSE is given in 'Text S8' and of the weights is given below

$$w_{a,\tau_1,t}^{m} = \frac{1}{(1+NRMSE_{a,\tau_1,t-1}^{m})}$$

where $w_{a,\tau_1,t-1}^{m}$ is the weight of serotype prediction during current prediction time $(t)$ for a given age group $(a)$, post-PCV period $(\tau_1)$, and model $(m)$, and $NRMSE_{a,\tau_1,t-1}^{m}$ is the normalised root mean square error (accuracy measure) of serotypes predictions during previous prediction time ($t$-1) as explained in the next section. Models with weights close to 1 contributed more to the ensemble predictions than models with weights close to 0.

**Supplementary Text 8**

*Calculation of the normalised root mean square error*

The normalised root mean square error (NRMSE) measured the accuracy of the model-predicted IPD case counts compared to the observed or reported IPD case counts during post-PCV period, and is given by

$$NRMSE_{\alpha,\tau_1,m} = \frac{\sqrt{\sum_{s=1}^{N}\left(I_{s,a,\tau_1} - I_{s,a,\tau_1}^{m}\right)^2 / N}}{Q_1\left(I_{s,a,\tau_1}\right) - Q_3\left(I_{s,a,\tau_1}\right)}$$

Where $NRMSE_{\alpha,s,\tau_1}$ is the normalised root mean square error between observations and predictions for a given age group $(a)$, post-PCV period $(\tau_1)$ and model $(m)$. $I_{s,a,\tau_1}$is the observed number of IPD cases for a given serotype $(s)$, age group $(a)$ and post-PCV period $(\tau_1)$. $I_{s,a,\tau_1}^{m}$ is the predicted number of IPD cases for the same serotype, age group and post-PCV period by a given model $(m)$, and $N$ is the total number of all considered non-vaccine serotypes. $Q_1$, $Q_3$ are 25$^{th}$, 75$^{th}$ quartiles (interquartile range), respectively, for normalizing the root mean square error so that this accuracy metric is comparable across age groups, PCV periods and models.

**Supplementary Text 9**

*Linear associations between observed and predicted post-PCV IPD cases*

We used Pearson correlation coefficient to quantify the relationship between observed and predicted post-PCV IPD cases as shown below.

$$Pearson_{\alpha,\tau_1} = \frac{\sum_{s=1}^{N}\left(I_{s,a,\tau_1} - \overline{I_{s,a,\tau_1}}\right)\left(I_{s,a,\tau_1}^{m} - \overline{I_{s,a,\tau_1}^{m}}\right)}{\sqrt{\sum_{s=1}^{N}\left(I_{s,a,\tau_1} - \overline{I_{s,a,\tau_1}}\right)^2 \sum_{s=1}^{N}\left(I_{s,a,\tau_1}^{m} - \overline{I_{s,a,\tau_1}^{m}}\right)^2}} ------ (6)$$

Where $I_{s,a,\tau_1}$ is the total number of observed IPD cases caused by serotype $(s)$ among individuals in age group $(\alpha)$ during post-PCV $(\tau_1)$, $I_{s,\tau_1}^{m}$ is similarly the total number of predicted IPD cases during post-PCV, $\overline{I_{s,a,\tau_1}}$ and $\overline{I_{s,a,\tau_1}^{m}}$ refer to the average number of observed and predicted IPD cases of each serotype $(s)$ among individuals in age group $(\alpha)$ during the post-PCV period, and $N$ is the total number of serotypes.

*Ranked association between observed and predicted post-PCV IPD cases*

We used Pearson correlation coefficient to quantify the relationship between observed and predicted post-PCV IPD cases as demonstrated below.

$$Spearman_{\alpha,\tau_1} = 1 - \frac{6 * \sum_{s} \Omega_{s,a,\tau_1}^{2}}{N(N^2 - 1)} ------ (7)$$

Where, $\Omega_s$ is the difference between paired ranks of observed and predicted IPD cases caused by serotype $(s)$ among individuals in age group $(\alpha)$ during post-PCV period, $N$ is the total number of all non-vaccine serotypes pairs between observed and predicted.

Supplementary Table 1. Relative effectiveness of pneumococcal conjugate vaccines against each serotype in carriage or in invasive pneumococcal disease (IPD) for ‘PCV7’ and ‘PCV13 relative to PCV7’.

| | Efficacy against carriage | | Efficacy against IPD | |
|---|---|---|---|---|
| Serotype | PCV7 | PCV13 | PCV7 | PCV13 |
| 1 | | 0.7 | | 0.87 |
| 3 | | 0.43 | | 0.80 |
| 4 | 0.63 | 0.48 | 0.93 | 0.97 |
| 5 | | 0.80 | | 0.87 |
| 6A* | | 0.34 | 0.94 | 0.86 |
| 6B* | 0.63 | 0.63 | 0.94 | 0.86 |
| 7F | | 0.92 | | 0.97 |
| 9V | 0.63 | 0.61 | 1.00 | 0.75 |
| 14 | 0.63 | 0.57 | 0.94 | 0.97 |
| 18C | 0.63 | 0.60 | 0.97 | 0.99 |
| 19A | | 0.29 | | 0.86 |
| 19F | 0.63 | 0.66 | 0.87 | 0.91 |
| 23F | 0.63 | 0.53 | 0.98 | 0.87 |
| NVT# | 0.00 | 0.00 | 0.00 | 0.00 |

PCV: Pneumococcal conjugate vaccines

PCV7: serotypes (4, [6A, 6B], 9V, 14, 18C, 19F, 23F)

PCV13: serotypes (1, 3, 4, 5, 6A, 6B, 7F, 9V, 14, 18C, 19A, 19F, 23F)

* [6A, 6B] - 6B assumed to cross protect 6A even though 6A is not part of PCV7

Estimates of relative effectiveness against carriage are from Wong et al. (2024) [9]

Estimates of relative effectiveness against IPD are from Ryman et al. (2024) [10]

### PCV7 or PCV13 is assumed to have a relative efficacy of 0 against any non-vaccine serotype (NVT) carriage or disease e.g., PCV has no effect on NVTs

Supplementary Table 2. Snapshot demonstration of the Ranking method and a few selected serotypes in a given age group, their pre-PCV share of carriage ('prevalence') and subsequent calculations of post-PCV serotype prevalence and the predicted change in serotype colonisation from pre- to post-PCV.

| | | $P_{s,a,\tau_0}$ | $R_0$ | $R_1$ | $P_{s,a,\tau_1} = \left(\rho_{s,a,\tau_0} \middle| R_1 = R_0\right)$ | $\varphi_s = \left(\frac{\rho_{s,a,\tau_1}}{\rho_{s,a,\tau_0}}\right)$ |
|---|---|---|---|---|---|---|
| Serotype | VT? | Pre-PCV prevalence | Pre-PCV prevalence rank | Post-PCV prevalence rank | Post-PCV prevalence | Expected change in serotype colonisation |
| 35B | No | 0.10 | 5 | 2 | 0.20 | 2 |
| 6B | Yes | 0.05 | 6 | | * | |
| 19F | Yes | 0.30 | 1 | | * | |
| 33F | No | 0.01 | 8 | 4 | 0.12 | 12 |
| 22F | No | 0.05 | 7 | 3 | 0.15 | 3 |
| 4 | Yes | 0.20 | 2 | | * | |
| 9V | Yes | 0.15 | 3 | | * | |
| 19A | No | 0.12 | 4 | 1 | 0.30 | 2.5 |

* post-PCV VT prevalence is determined by: $P_{s,a,\tau_1} = \rho_{s,a,\tau_0} * (1 - \zeta_s)$, where $\zeta_s$ is the relative efficacy of PCV against carriage of each serotype $s$

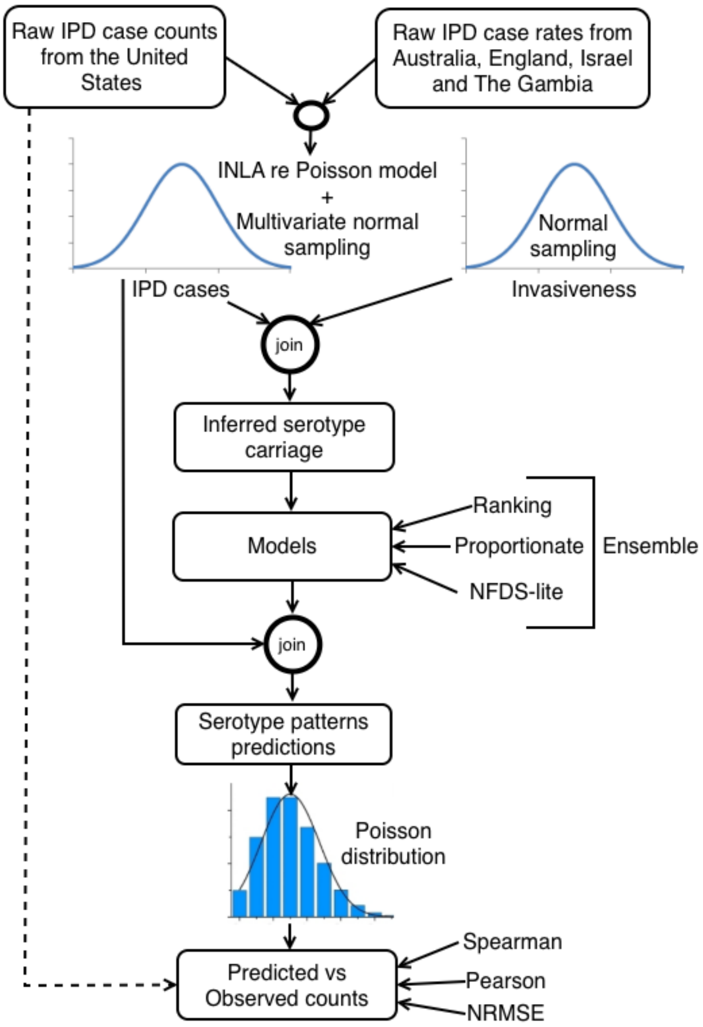


Supplementary Figure 1. Modeling overview of forecasting invasive pneumococcal disease (IPD) case frequency caused by non-vaccine serotypes. The flow diagram shows different steps in prediction modelling; starting with augmenting raw IPD case counts data in the United States and from other countries (during pre-PCV base periods). Then, inferring serotype carriage from serotype-specific invasiveness and base serotype-specific IPD cases. Then, using inferred carriage, by different models, to predict the expected change in serotype colonisation during post-PCV and combining it with base serotype-specific IPD estimates to predict post-PCV IPD cases. Lastly, taking a Poisson sample of the 10,000 IPD predictions per serotype and comparing it to the original IPD case data reported from the US.

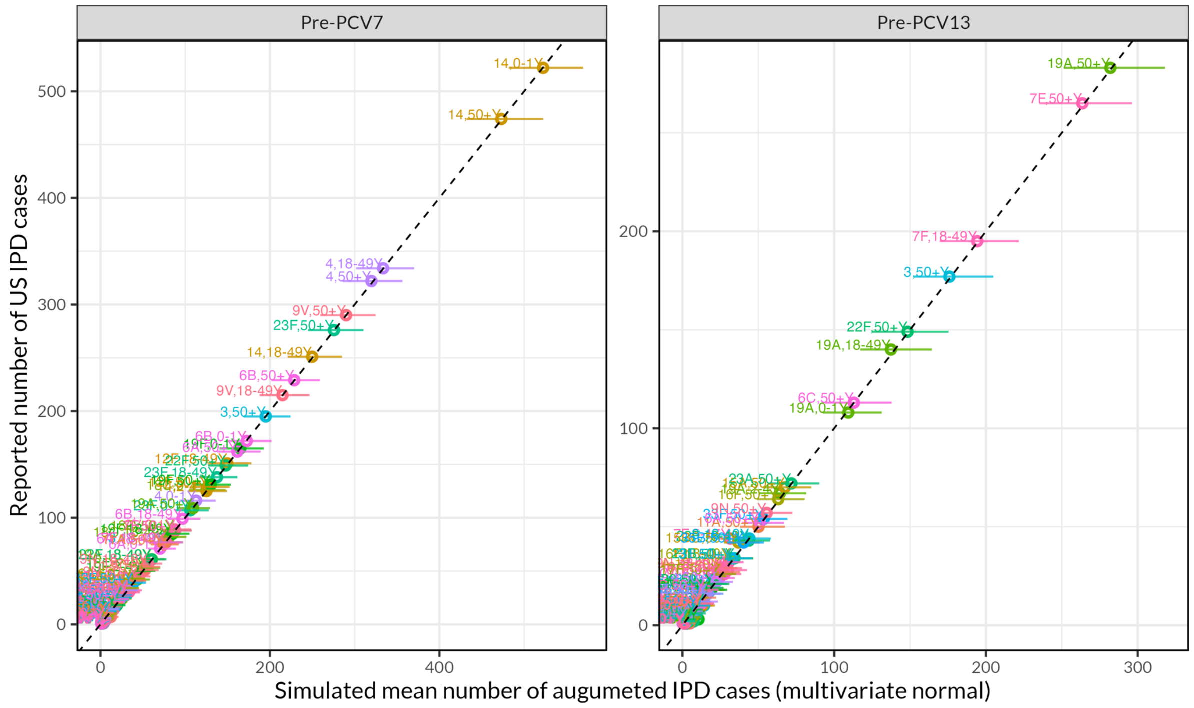


Supplementary Figure 2. Observed vs predicted IPD cases using a random effects Poisson regression model stratified by PCV periods. The Poisson model was fitted in INLA to each age group and PCV period, separately. The x-axis represents the simulated mean of 10,000 samples of fitted values and the y-axis represents the reported or observed IPD cases in the United States. Overall, the plot shows very high correlation between observed and simulated mean values. The pre-PCV ('pre-PCV7' and 'pre-PCV13') predictions and their uncertainty are used in downstream analyses to forecast post-PCV serotype patterns.

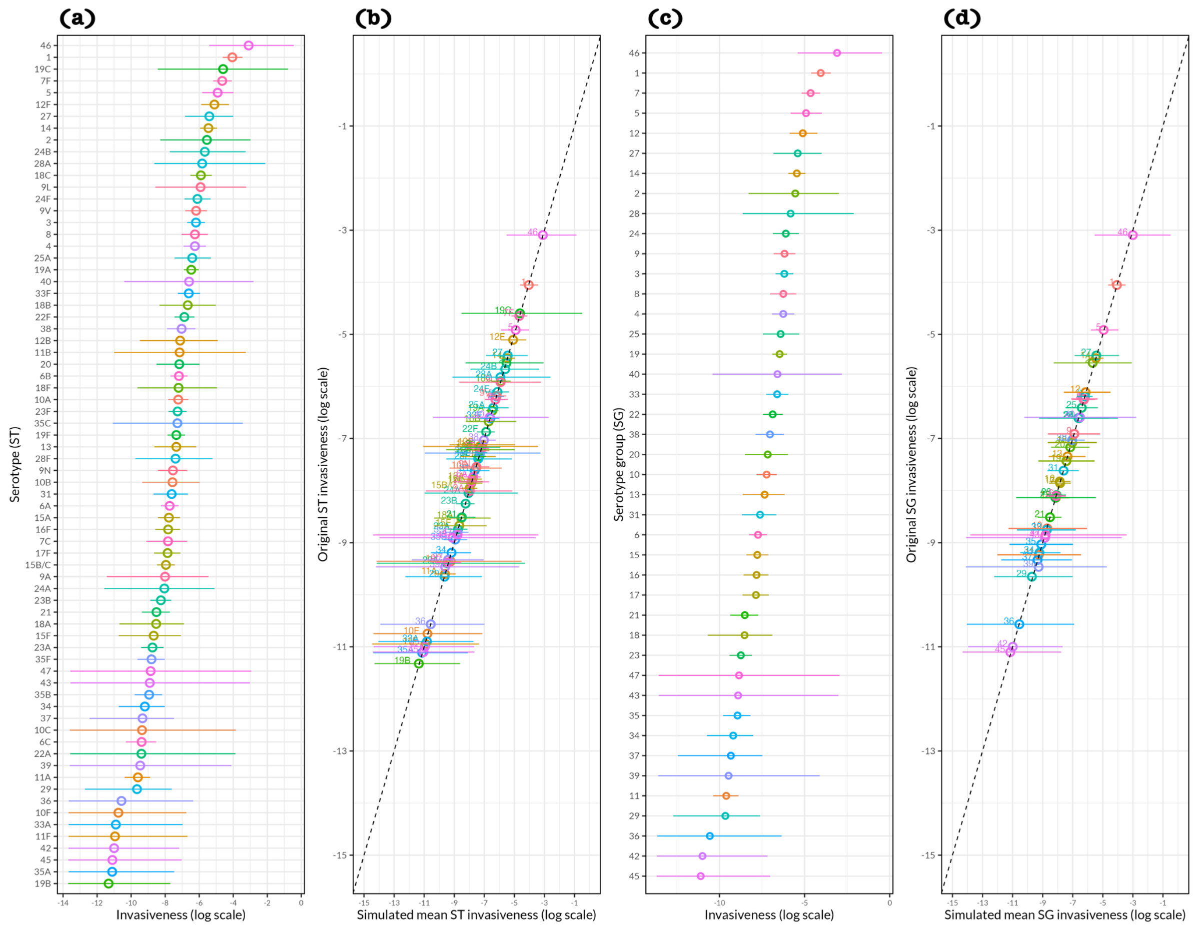


Supplementary Figure 3. Distribution of serotype, serogroup and their invasiveness, and the association between estimated (original) serotype or serogroup invasiveness and simulated mean serotype or serogroup invasiveness. In all plots, a circle point is a point estimate and horizontal line through the circle point is the 95% confidence interval of a point estimate. (A) Invasiveness of each serotype from Løchen et al. [7] . (B) Normal distribution-simulated mean invasiveness of each serotype compared to original serotype invasiveness in panel A, and details of simulation are explained in Supplementary Text 2. (C) Invasiveness of each original serogroup calculated as a mean of invasiveness of serotypes within serogroup as explained in Supplementary Text 2. (D) Normal distribution-simulated mean invasiveness of each serogroup compared to estimated serogroup invasiveness from panel C, and details of simulation are explained in Supplementary Text 2. Overall, the plots show very high correlation between original and simulated mean invasiveness of serotypes and serogroups. The simulated invasiveness of serotypes and serogroups are used in downstream analyses.

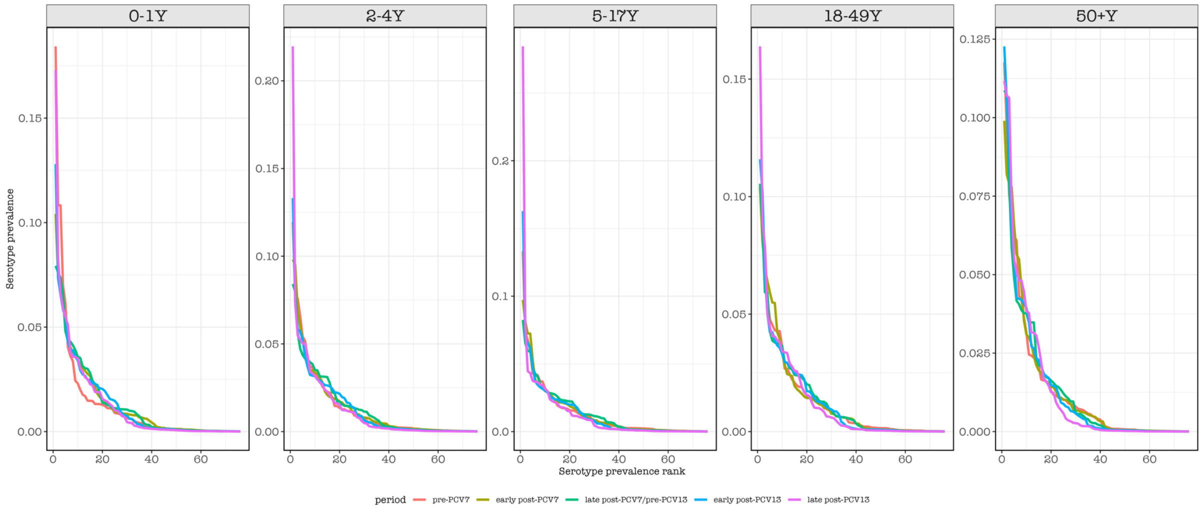


Supplementary Figure 4. Serotype prevalence ranks stratified by age groups and PCV periods in the United States. The plot shows stability in serotype prevalence ranking overtime where the y-axis is the prevalence of each serotype in total carriage, the x-axis is the rank order assigned to each serotype within the age group (0-1y, 2-4y, 5-17y, 18-49y, and ≥50y) and wihtin the PCV period (pre-PCV, 1998-1999; early post-PCV7, 2000-2006, late post-PCV7 or pre-PCV13, 2007-2009; early post-PCV13, 2010-2014 and late post-PCV13, 2015-2019).

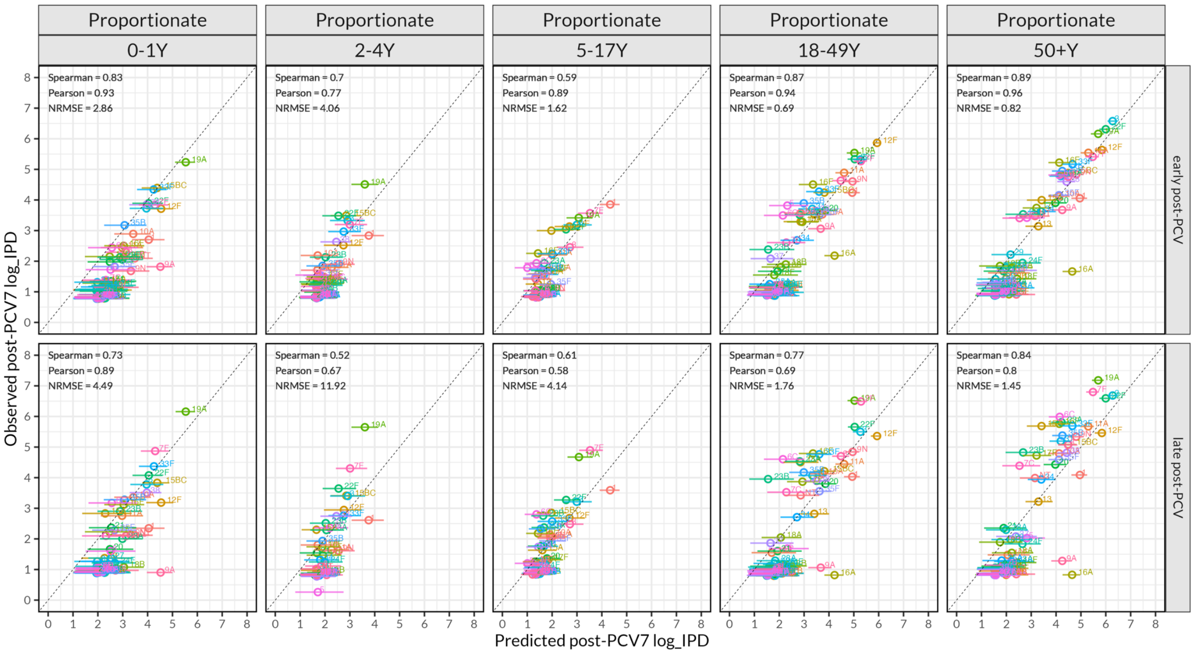


Supplementary Figure 5. Comparison of observed and predicted post-PCV7 invasive pneumococcal disease (IPD) frequency by age group and post-vaccination period based on **Proportionate model**. This figure illustrates the relationship between observed and model-predicted post-PCV7 IPD frequency (log scale) across pneumococcal serotypes, stratified by five age groups (0-1y, 2-4y, 5-17y, 18-49y, and ≥50y,) and two post-vaccine periods (rows, early post-PCV7 and late post-PCV7 defined for 2000-2004 and 2005-2009, respectively). Each point represents an individual pneumococcal serotype. The dashed diagonal line denotes the 1:1 relationship, representing perfect agreement between observed and predicted values. Model performance within each panel is summarized using three statistical metrics: Spearman correlation coefficient, Pearson correlation coefficient, and normalized root mean square error (NRMSE). Higher correlation values and lower NRMSE indicate better predictive accuracy.

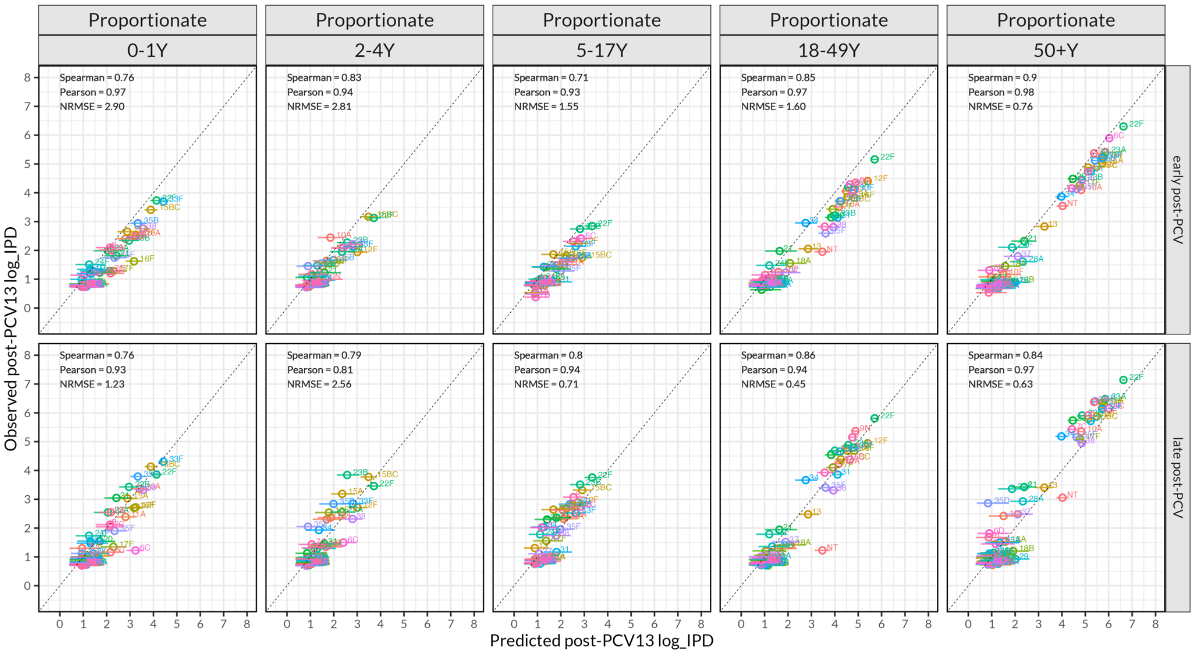


Supplementary Figure 6. Comparison of observed and predicted post-PCV13 invasive pneumococcal disease (IPD) frequency by age group and post-vaccination period based on **Proportionate model**. This figure illustrates the relationship between observed and model-predicted post-PCV13 IPD frequency (log scale) across pneumococcal serotypes, stratified by five age groups (0-1y, 2-4y, 5-17y, 18-49y, and ≥50y,) and two post-vaccine periods (early post-PCV13 and late post-PCV13 defined for 2010-2014 and 2015-2019, respectively). Each point represents an individual pneumococcal serotype. The dashed diagonal line denotes the 1:1 relationship, representing perfect agreement between observed and predicted values. Model performance within each panel is summarized using three statistical metrics: Spearman correlation coefficient, Pearson correlation coefficient, and normalized root mean square error (NRMSE). Higher correlation values and lower NRMSE indicate better predictive accuracy.

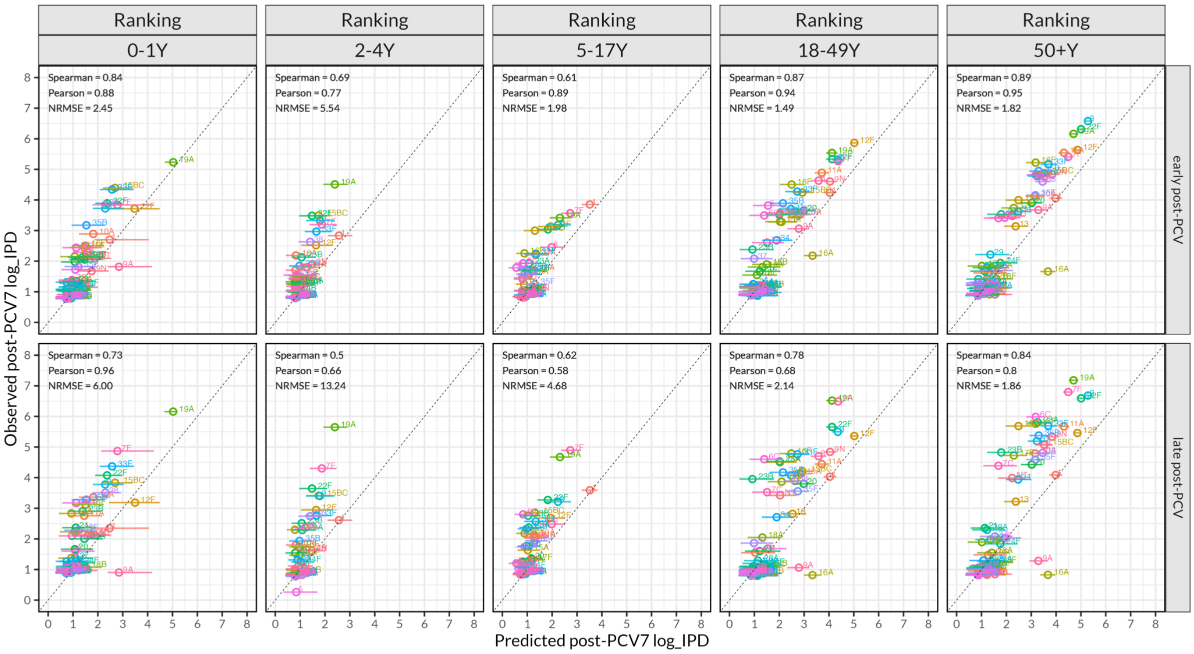


Supplementary Figure 7. Comparison of observed and predicted post-PCV7 invasive pneumococcal disease (IPD) frequency by age group and post-vaccination period based on **Ranking model**. This figure illustrates the relationship between observed and model-predicted post-PCV7 IPD frequency (log scale) across pneumococcal serotypes, stratified by five age groups (0-1y, 2-4y, 5-17y, 18-49y, and ≥50y) and two post-vaccine periods (early post-PCV7 and late post-PCV7 defined for 2000-2004 and 2005-2009, respectively). Each point represents an individual pneumococcal serotype. The dashed diagonal line denotes the 1:1 relationship, representing perfect agreement between observed and predicted values. Model performance within each panel is summarized using three statistical metrics: Spearman correlation coefficient, Pearson correlation coefficient, and normalized root mean square error (NRMSE). Higher correlation values and lower NRMSE indicate better predictive accuracy.

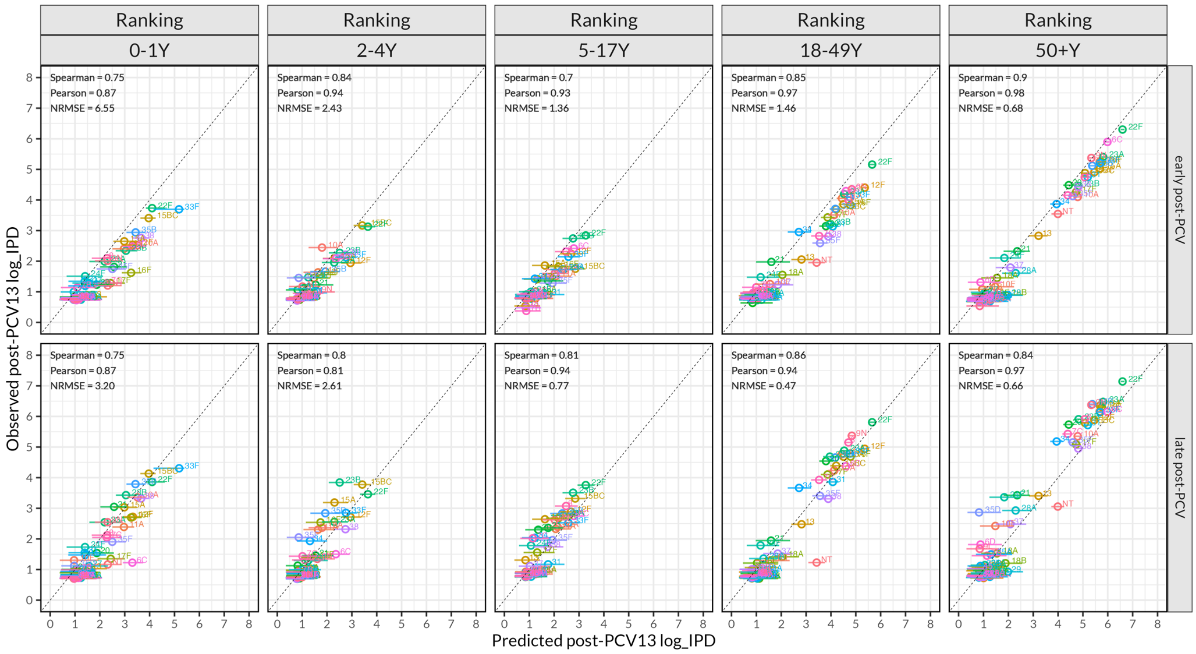


Supplementary Figure 8. Comparison of observed and predicted post-PCV13 invasive pneumococcal disease (IPD) frequency by age group and post-vaccination period based on **Ranking model**. This figure illustrates the relationship between observed and model-predicted post-PCV13 IPD frequency (log scale) across pneumococcal serotypes, stratified by five age groups (0-1y, 2-4y, 5-17y, 18-49y, and ≥50y,) and two post-vaccine periods (rows, early post-PCV13 and late post-PCV13 defined for 2010-2014 and 2015-2019, respectively). Each point represents an individual pneumococcal serotype. The dashed diagonal line denotes the 1:1 relationship, representing perfect agreement between observed and predicted values. Model performance within each panel is summarized using three statistical metrics: Spearman correlation coefficient, Pearson correlation coefficient, and normalized root mean square error (NRMSE). Higher correlation values and lower NRMSE indicate better predictive accuracy.

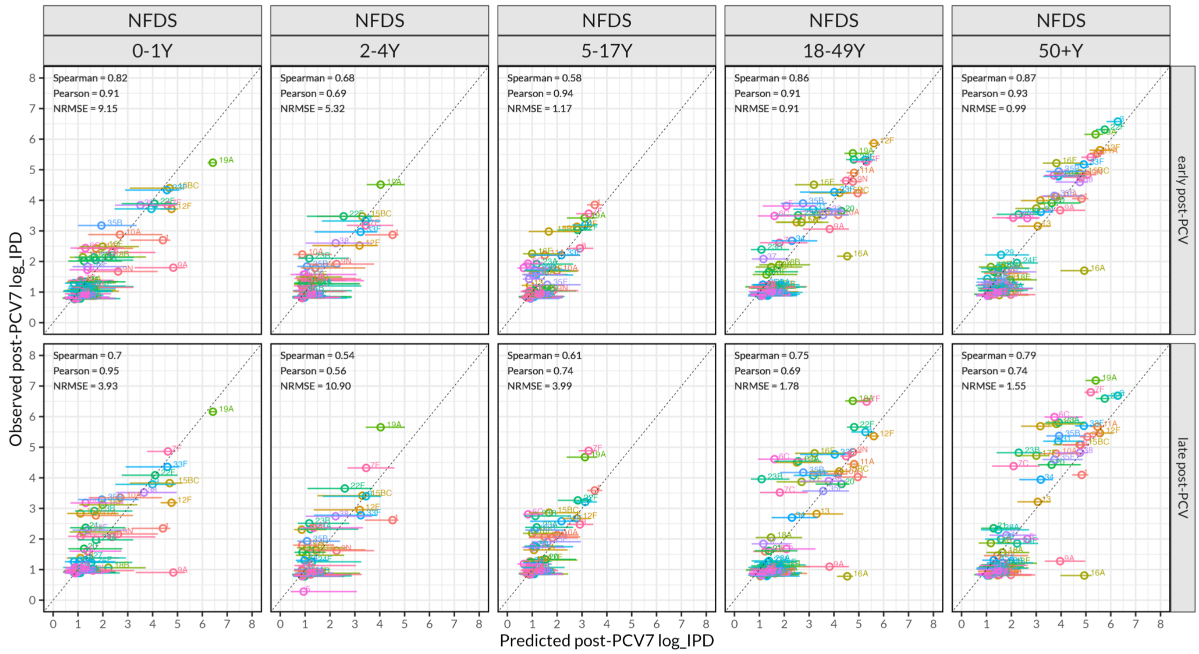


Supplementary Figure 9. Comparison of observed and predicted post-PCV7 invasive pneumococcal disease (IPD) frequency by age group and post-vaccination period based on **Negative Frequency-Dependent Selection lite (NFDS-lite) model**. This figure illustrates the relationship between observed and model-predicted post-PCV7 IPD frequency (log scale) across pneumococcal serotypes, stratified by five age groups (0-1y, 2-4y, 5-17y, 18-49y, and ≥50y,) and two post-vaccine periods (early post-PCV7 and late post-PCV7 defined for 2000-2004 and 2005-2009, respectively). Each point represents an individual pneumococcal serotype. The dashed diagonal line denotes the 1:1 relationship, representing perfect agreement between observed and predicted values. Model performance within each panel is summarized using three statistical metrics: Spearman correlation coefficient, Pearson correlation coefficient, and normalized root mean square error (NRMSE). Higher correlation values and lower NRMSE indicate better predictive accuracy.

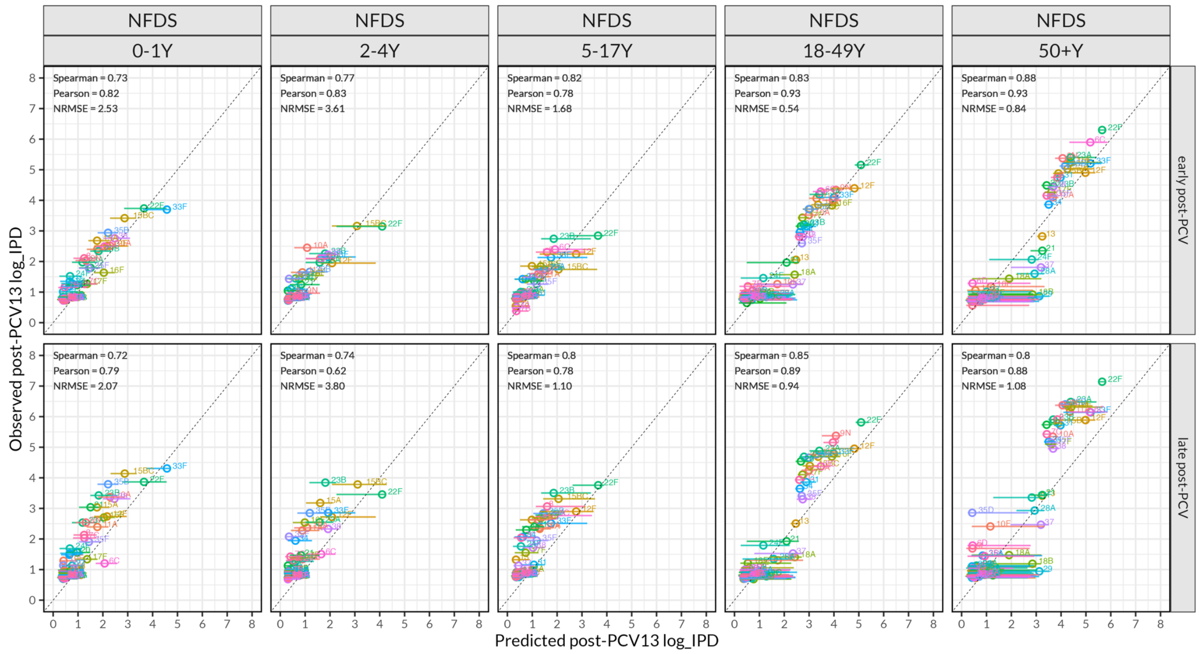


Supplementary Figure 10. Comparison of observed and predicted post-PCV13 invasive pneumococcal disease (IPD) frequency by age group and post-vaccination period based on **Negative Frequency-Dependent Selection (NFDS-lite) model**. This figure illustrates the relationship between observed and model-predicted post-PCV13 IPD frequency (log scale) across pneumococcal serotypes, stratified by five age groups (0-1y, 2-4y, 5-17y, 18-49y, and ≥50y,) and two post-vaccine periods (early post-PCV13 and late post-PCV13 defined for 2010-2014 and 2015-2019, respectively). Each point represents an individual pneumococcal serotype. The dashed diagonal line denotes the 1:1 relationship, representing perfect agreement between observed and predicted values. Model performance within each panel is summarized using three statistical metrics: Spearman correlation coefficient, Pearson correlation coefficient, and normalized root mean square error (NRMSE). Higher correlation values and lower NRMSE indicate better predictive accuracy.

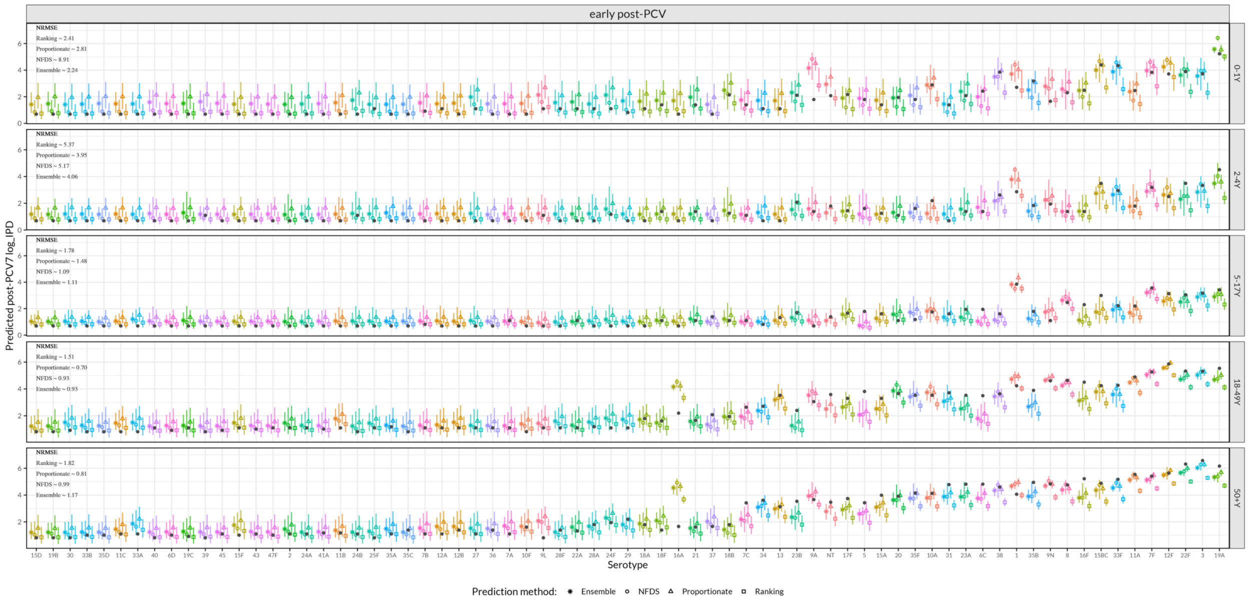


Supplementary Figure 11. The predictions of the **early post-PCV7** serotype invasive pneumococcal disease (IPD) cases (log scale) stratified by five age groups (0-1y, 2-4y, 5-17y, 18-49y, and 50+y). The x-axis represents individual serotype and y-axis represents model-based predictions of the number of IPD cases in the early post-PCV7 period. Predictions were generated using three modeling approaches and Ensemble: Ranking (square), Proportionate (triangle), Negative frequency-dependent selection variant (NFDS-lite) (circle), and Ensemble (asterisk). Each point shape represents the predicted case count estimate and the vertical line through it is the corresponding 95% confidence interval. The black dots in the plot represent reported IPD case count in the United States. The legend within each panel summarizes models' performances based using the normalized root mean square error (NRMSE).

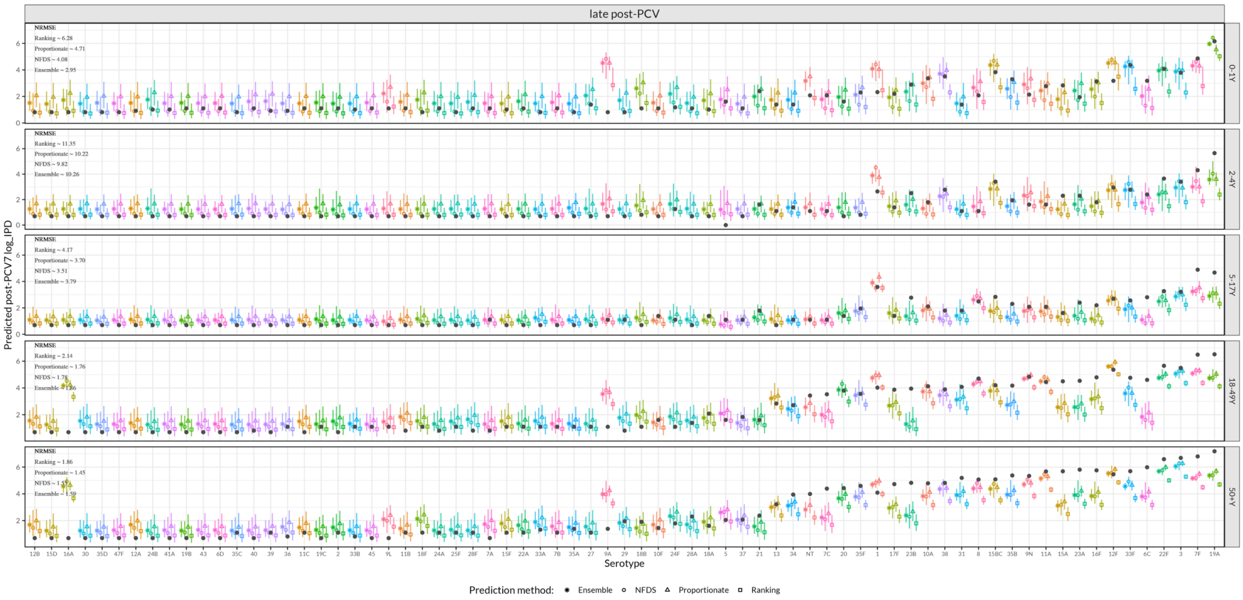


Supplementary Figure 12. The predictions of **late post-PCV7** serotype invasive pneumococcal disease (IPD) cases (log scale) stratified by five age groups (0-1y, 2-4y, 5-17y, 18-49y, and 50+y). The x-axis represents individual serotype and y-axis represents model-based predictions of the number of IPD cases in the late post-PCV7 period. Predictions were generated using three modeling approaches and Ensemble: Ranking (square), Proportionate (triangle), Negative frequency-dependent selection variant (NFDS-lite) (circle), and Ensemble (asterisk). Each point shape represents the predicted case count estimate and the vertical line through it is the corresponding 95% confidence interval. The black dots in the plot represent reported IPD case count in the United States The legend within each panel summarizes models' performances using the normalized root mean square error (NRMSE).

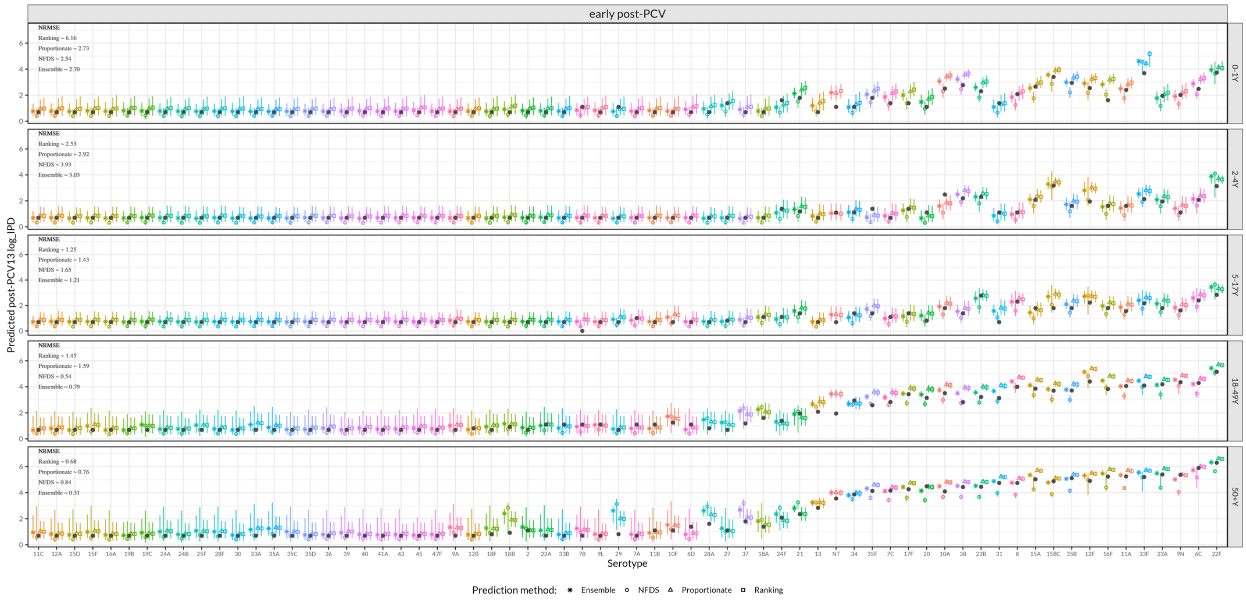


Supplementary Figure 13. The predictions of **early post-PCV13** serotype invasive pneumococcal disease (IPD) cases (log scale) stratified by five age groups (0-1y, 2-4y, 5-17y, 18-49y, and 50+y). The x-axis represents individual serotype and y-axis represents model-based predictions of the number of IPD cases in the early post-PCV13 period. Predictions were generated using three modeling approaches and Ensemble: Ranking (square), Proportionate (triangle), Negative frequency-dependent selection variant (NFDS-lite) (circle), and Ensemble (asterisk). Each point shape represents the predicted case count estimate and the vertical line through it is the corresponding 95% confidence interval. The black dots in the plot represent reported IPD case count in the United States The legend within each panel summarizes models' performances using the normalized root mean square error (NRMSE).

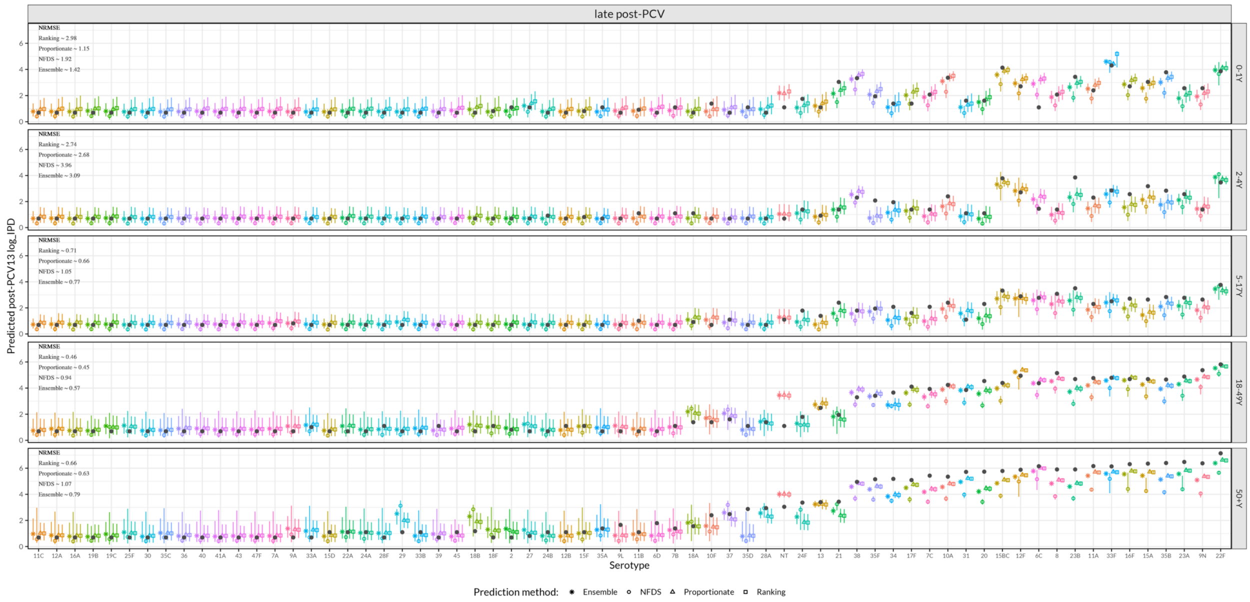


Supplementary Figure 14. The predictions of **late post-PCV13** serotype invasive pneumococcal disease (IPD) cases (log scale) stratified by five age groups (0-1y, 2-4y, 5-17y, 18-49y, and 50+y). The x-axis represents individual serotype and y-axis represents model-based predictions of the number of IPD cases in the late post-PCV13 period. Predictions were generated using three modeling approaches and Ensemble: Ranking (square), Proportionate (triangle), Negative frequency-dependent selection variant (NFDS-lite) (circle), and Ensemble (asterisk). Each point shape represents the predicted case count estimate and the vertical line through it is the corresponding 95% confidence interval. The black dots in the plot represent reported IPD case count in the United States The legend within each panel summarizes models' performances using the normalized root mean square error (NRMSE).

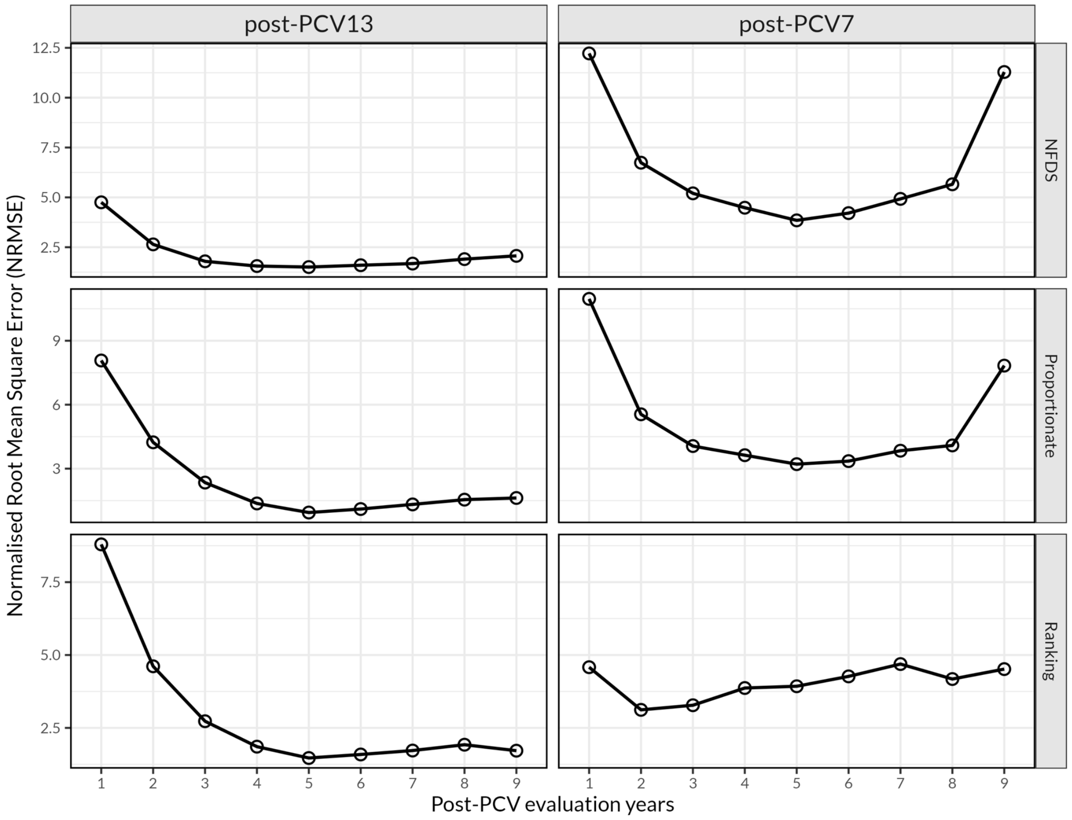


Supplementary Figure 15. Model performance during different post-PCV evaluation periods, by prediction method. This figure shows disparities in model performance for predicting post-PCV7 and post-PCV13 invasive pneumococcal disease (IPD) cases over different periods (indicated by circles) using the three modeling approaches (NFDS-lite, Proportionate, and Ranking). The y-axis represents the normalized root mean square error (NRMSE), a measure of prediction accuracy where lower values indicate better predictions. The x-axis corresponds to post-PCV7 evaluation years from 1 through 9 following PCV introduction. The figure shows that the optimal number of post-vaccine years for predicting post-PCV7 and post-PCV13 serotype patterns is largely between 4 and 6 years, irrespective of prediction methods.

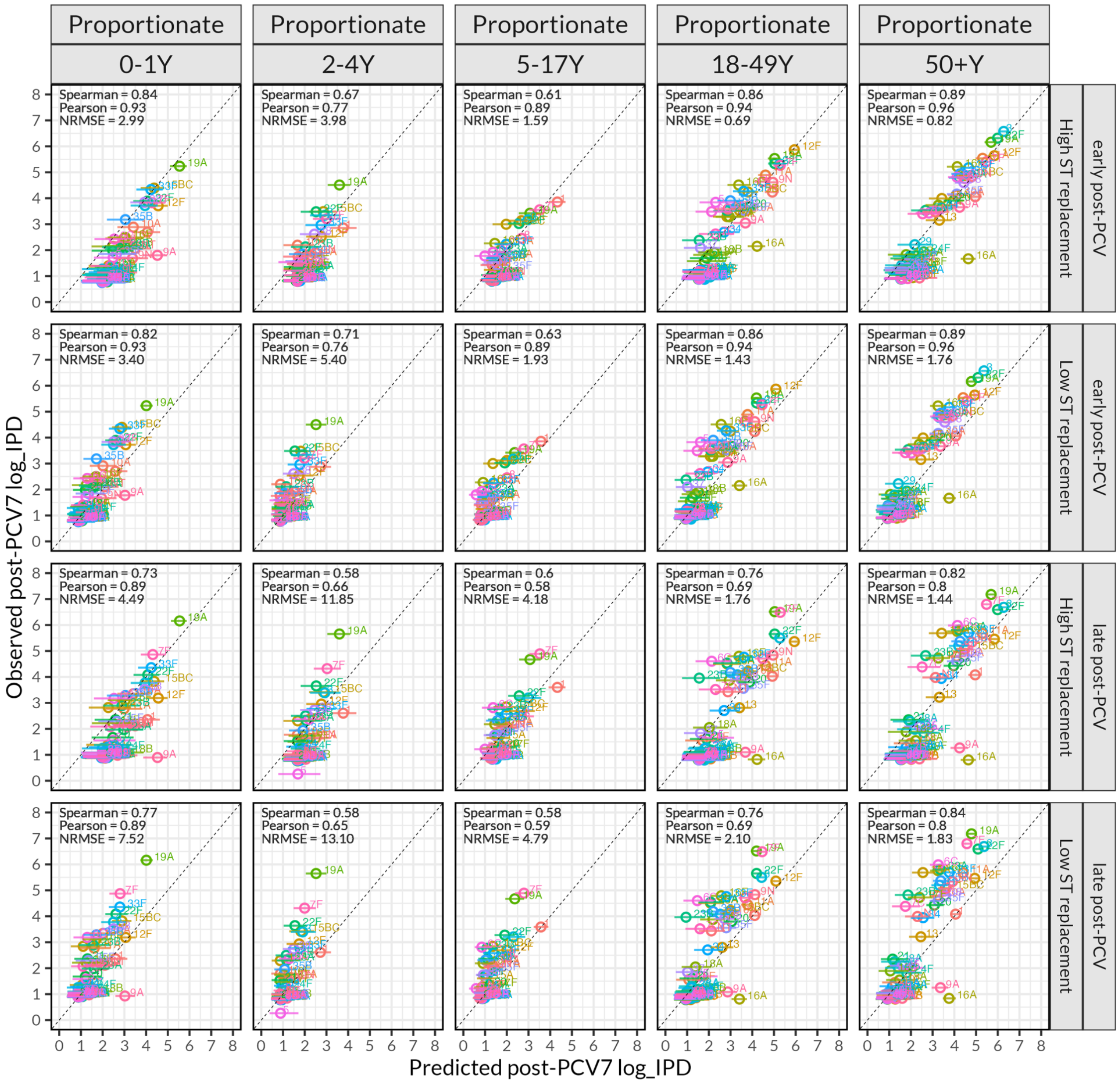


Supplementary Figure 16. Comparison of predictions from the proportionate model based on low proportion of replaced serotypes of 5% compared to 95% (high) used in the main findings. The plot compares observed and predicted post-PCV7 IPD frequency (log scale) across serotypes, stratified by five age groups, post-vaccine periods (early and late post-PCV7 defined for 2000-2004 and 2005-2009) and low/high serotype replacement. Each point represents an individual serotype with dashed diagonal line denoting a perfect agreement. Model performance within each panel is summarized using Spearman and Pearson correlation coefficients, and normalized root mean square error (NRMSE). Higher correlation values and lower NRMSE indicate better predictive accuracy.

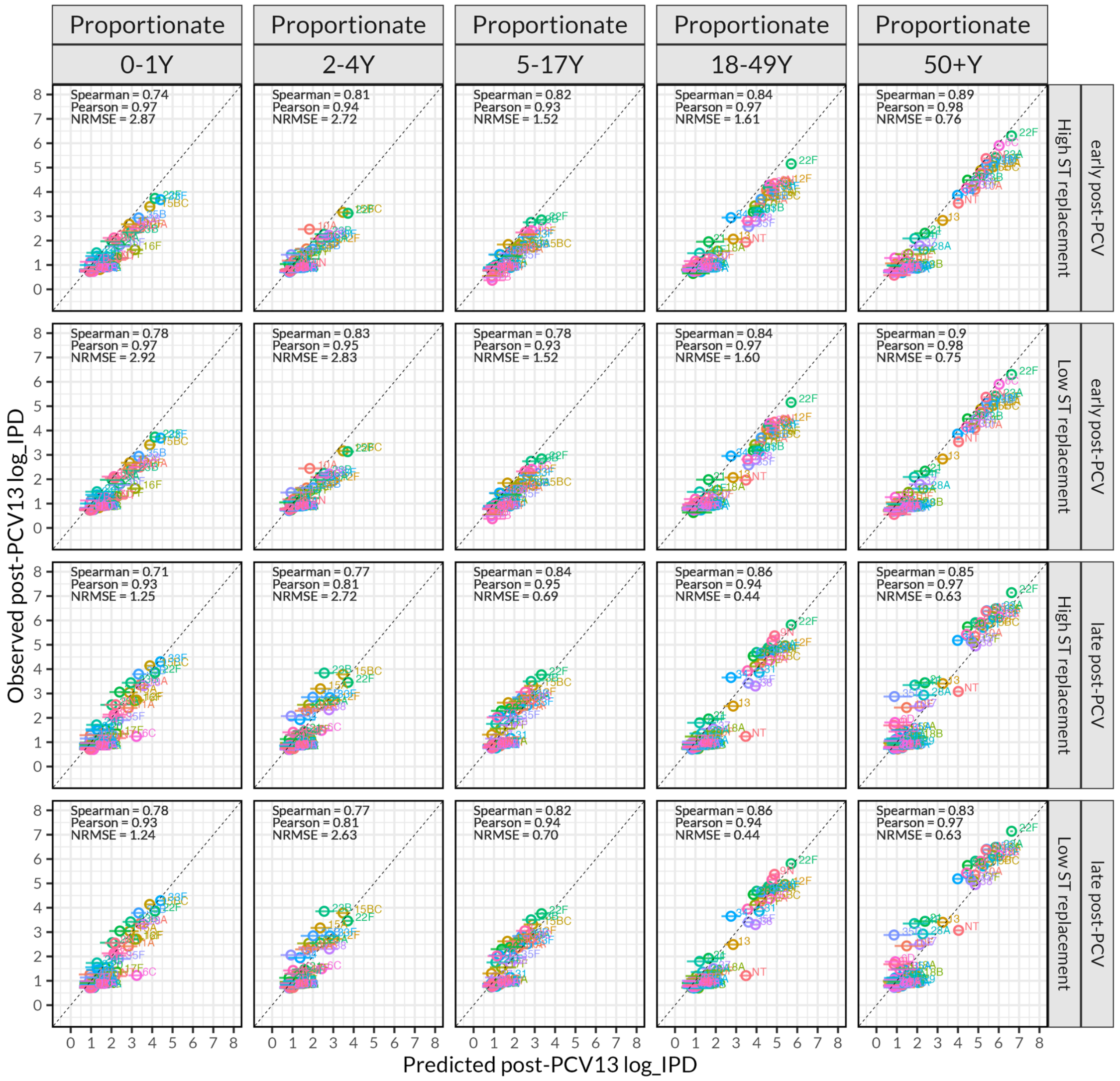


Supplementary Figure 17. Comparison of predictions from the proportionate model based on low proportion of replaced serotypes of 5% compared to 95% (high) used in the main findings. The plot compares observed and predicted post-PCV13 IPD frequency (log scale) across serotypes, stratified by five age groups, post-vaccine periods (early and late post-PCV13 defined for 2010-2014 and 2015-2019) and low/high serotype replacement. Each point represents an individual serotype with dashed diagonal line denoting a perfect agreement. Model performance within each panel is summarized using Spearman and Pearson correlation coefficients, and normalized root mean square error (NRMSE). Higher correlation values and lower NRMSE indicate better predictive accuracy.
